# Genome-wide Association Study of Complex Lipid Species: Results from the Population-based Rhineland Study

**DOI:** 10.1101/2024.12.04.24318368

**Authors:** Elvire N. Landstra, Mohammed A. Imtiaz, Valentina Talevi, Fabian Eichelmann, Matthias B. Schulze, N. Ahmad Aziz, Monique M.B. Breteler

**Affiliations:** Population Health Sciences, German Centre for Neurodegenerative Diseases (DZNE), Bonn, Germany; Department of Molecular Epidemiology, German Institute of Human Nutrition Potsdam-Rehbruecke, Nuthetal, Germany; German Center for Diabetes Research (DZD), Neuherberg, Germany; Institute of Nutritional Science, University of Potsdam, Nuthetal, Germany; Department of Neurology, Faculty of Medicine, University of Bonn, Bonn, Germany; Institute for Medical Biometry, Informatics and Epidemiology (IMBIE), Faculty of Medicine, University of Bonn, Germany

**Keywords:** genome-wide association study, complex lipids, lipidome, ceramides, triacylglycerols, sphingolipids, lipid composition, lipid saturation, expression quantitative trait loci

## Abstract

The human lipidome comprises numerous complex lipids, dysregulation of which can contribute to the pathogenesis of a wide range of diseases. Despite the high heritability of parts of the lipidome, the genetic architecture of many circulating lipid species and their structure remains mostly unknown. Thus, we performed genome-wide association studies on 970 lipid species and 267 fatty acid composite measures using samples from the population-based Rhineland Study (n=6,096). We validated our findings using corresponding data from two other independent cohorts, including FinnGen (n=7,266) and EPIC-Potsdam (n=1,188). Out of 217 lead genomic loci, we found 135 to be novel, such as *FDFT1*. Using mendelian randomization and individual-level gene expression data, we identified five possible causal associations between candidate genes and corresponding lipid species, including *FDFT1*-diacylglycerol (16:0/18:0). Our findings provide new insights into the intricate genetic underpinnings of lipid metabolism, which may facilitate risk stratification and discovery of new therapeutic targets.

## Introduction

The metabolome – the collection of circulating metabolites – exhibits substantial inter-individual variability that may partly reflect differences in individual susceptibility to a variety of diseases, including cardio-metabolic and neurodegenerative disorders (1–3). In particular, circulating levels of complex lipids, such as ceramides (CERs) and triacylglycerols (TAGs), are increasingly recognized as important correlates of age-related structural and functional changes in a variety of organs and tissues, including the brain, heart, kidney, and fat and muscle tissue (4–13). Importantly, circulating complex lipid levels have also been associated with a range of common cardiovascular, metabolic and neurodegenerative diseases (14–16).

A large degree of the variation in the levels of circulating metabolites, including lipids, is thought to be genetically determined (17–19). Indeed, previous genome-wide association studies (GWASs) have identified several genetic variants linked to circulating levels of certain triglycerides, phospholipids and sphingolipids (17,18,20,21). These prior findings highlighted the importance of distinguishing between different lipid species rather than classes, as the genetic determinants of circulating lipid levels depended on their specific biochemical properties, such as the lipid species’ carbon chain length and degree of saturation, rather than on their specific class (2,19). However, with more than 26,000 lipids curated to date, much of their properties still remain to be elucidated (22), including the genetic basis of the majority of complex lipid species, as well as their full fatty acid (FA) composition (17,18). Indeed, the largest study to date employed a panel containing only 596 lipid species across 33 classes (23). Therefore, there are a large number of lipid classes whose genetic basis has not been investigated at all. Furthermore, even for those lipids included in previous GWAS studies, the genetic basis of their FA composition within and across classes was not systematically assessed.

As complex lipids have been critically implicated in the pathogenesis of many cardiometabolic and neurodegenerative diseases (14–16), the identification of their genetic determinants could improve our understanding of the molecular mechanisms underlying these disorders and yield potential new therapeutic targets. Therefore, taking advantage of recent technological innovations that enable accurate high-throughput profiling of a large number of lipid species that previously could not be measured at scale, we aimed to further disentangle the genetic architecture of complex lipid species and fatty acid composition, primarily focusing on those metabolites whose genetic determinants were so far unknown. We used data from the Rhineland Study, an ongoing population-based cohort study in Bonn, Germany, to perform a GWAS on circulating complex lipid levels and composition measured using the Metabolon Complex Lipids Platform. Importantly, we could validate our findings using corresponding data from two other independent population-based cohorts, namely the FinnGen and EPIC-Potsdam studies (24,25). Subsequently, we contextualized our results through heritability estimates, phenome-wide association studies (PheWAS), and genetic correlation and mendelian randomization (MR) analyses. Finally, we also functionally validated several candidate genes using concomitantly available individual-level gene expression data. Our systematic approach uncovered a large number of novel genetic variants, metabolic quantitative trait loci (mQTLs), as well as candidate genes and pathways involved in complex lipid metabolism and FA composition.

## Results

### Sample characteristics

We included Rhineland Study participants who enrolled before the 26^th^ of November 2021 (n= 8,318), and who had complete lipidomic and genetic data available (n = 6,096). The mean age was 55.9 years (range: 30-95) and 56.3% were women (**Table 1**). Comparing the included participants to those who were excluded because of missing data in any of the variables, we observed higher high-density lipoprotein cholesterol (HDL-C) (p-value < 0.001) and lower triglyceride levels (p-value < 0.001) (**Table 1**). An overview of the total lipid class concentrations can be found in **Supplementary Table 1**.

**Table 1.** Overview of the Rhineland Study sample characteristics comparing included versus excluded.

| Characteristic | Included participants<br>(n= 6,096) | Excluded participants<br>(n= 2,222) | P-value* |
| --- | --- | --- | --- |
| Age (years) | 55.9 (13.7) | 55.8 (14.3) | 0.762 |
| Sex (women) | 3435 (56.3) | 1259 (56.7) | 0.806 |
| LDL-C (mg/dL) | 125.7 (35.6) | 124.3 (35.1) | 0.153 |
| HDL-C (mg/dL) | 63.0 (17.6) | 60.6 (17.3) | **<0.001** |
| Triglycerides (mg/dL) | 109.2 (64.4) | 117.0 (84.1) | **<0.001** |
| Cholesterol (mg/dL) | 199.2 (39.3) | 198.0 (39.0) | 0.315 |
| Use of lipid-lowering medication (yes) | 721 (12.0) | 253 (11.7) | 0.727 |

Data displayed represent the number of participants (percentages) for categorical variables and the mean (standard deviation) for continuous variables. Abbreviations: HDL-C = high-density lipoprotein cholesterol, LDL-C = low-density lipoprotein cholesterol.
\*Adjusted for age and sex.

### Replication cohorts

We used complete data from the FinnGenn cohort (n =7,266) and a subsample of the EPIC-Potsdam cohort (n = 1,188) to replicate our findings. FinnGen and EPIC-Potsdam participants had an average age of 55.8 and 50.3 years and consisted of 64.7 % and 60.3% women, respectively (**Supplementary Table 2**).

### GWAS of complex lipid species

We first ran a GWAS on lipid species adjusting for age, sex and the first 10 genetic principal components (model 1). Lipid species were quantified on the Metabolon Complex Lipids Platform, which identified 970 species across 14 lipid classes, where the number of species per class ranged between 12 and 518. Lipid species were characterized by a complete identification of the total number of carbons and double bonds and at least one FA tail (e.g., diacylglycerols DAG(16:1/20:0), ceramides (CER) 20:1, and triacylglycerols (TAG) 53:5(FA18:3)) (**Supplementary Table 3**).

We identified a total of 57 genomic loci associated with the levels of different lipid species after adjustment for sex, age and the first 10 genetic principal components. These genomic loci were identified by merging the linkage disequilibrium (LD) blocks of 1296 metabolome-wide (p < 2.27E-10) independent significant SNPs (using r^2^ ≥ 0.6 for defining independent significant SNPs) within 250 kb of each other, and 218 lead SNPs (using r^2^ ≥ 0.1 for the clumping of independent significant SNPs). Out of the 57 identified loci, 30 were novel across all lipid categories and classes (**Figure 1**). Among these novel loci were *NIPAL1*, *ABCA7*, *GM2A*, *FDFT1, ITGA11*, which were most strongly associated with hexosylceramide (HCER) (FA22:1), lactosylceramide (LCER) (FA24:1), LCER(FA20:1)), DAG(16:0/18:0)) and phosphatidylethanolamines (PE) (18:0/18:3), respectively. The identified loci with the highest statistical significance were found on chromosome 11 (*FADS2*, *OR4C15*, *TRIM48*), followed by loci on chromosome 6 (*ELOVL2*, *GABBR1*). Although many genomic loci were unique to different lipid categories, some were also shared, particularly between neutral lipids and sphingolipids (**Figure 1**). Specifically, we found lipidome-wide significant SNPs for 638 out of 970 lipid species. The lipid species within the sphingomyelins (SMs) (100%), TAGs (78.2%) and DAGs (75.9%) classes had the highest number of genetic associations. On the other hand, the lipid species within the phosphatidylethanolamines ethers (PEOs) (30%) class had only a few genetic associations. The associated genomic loci and their independent significant SNPs identified in the GWAS on lipid species can be found in the **Supplementary Data 1**.

**Figure 1:**
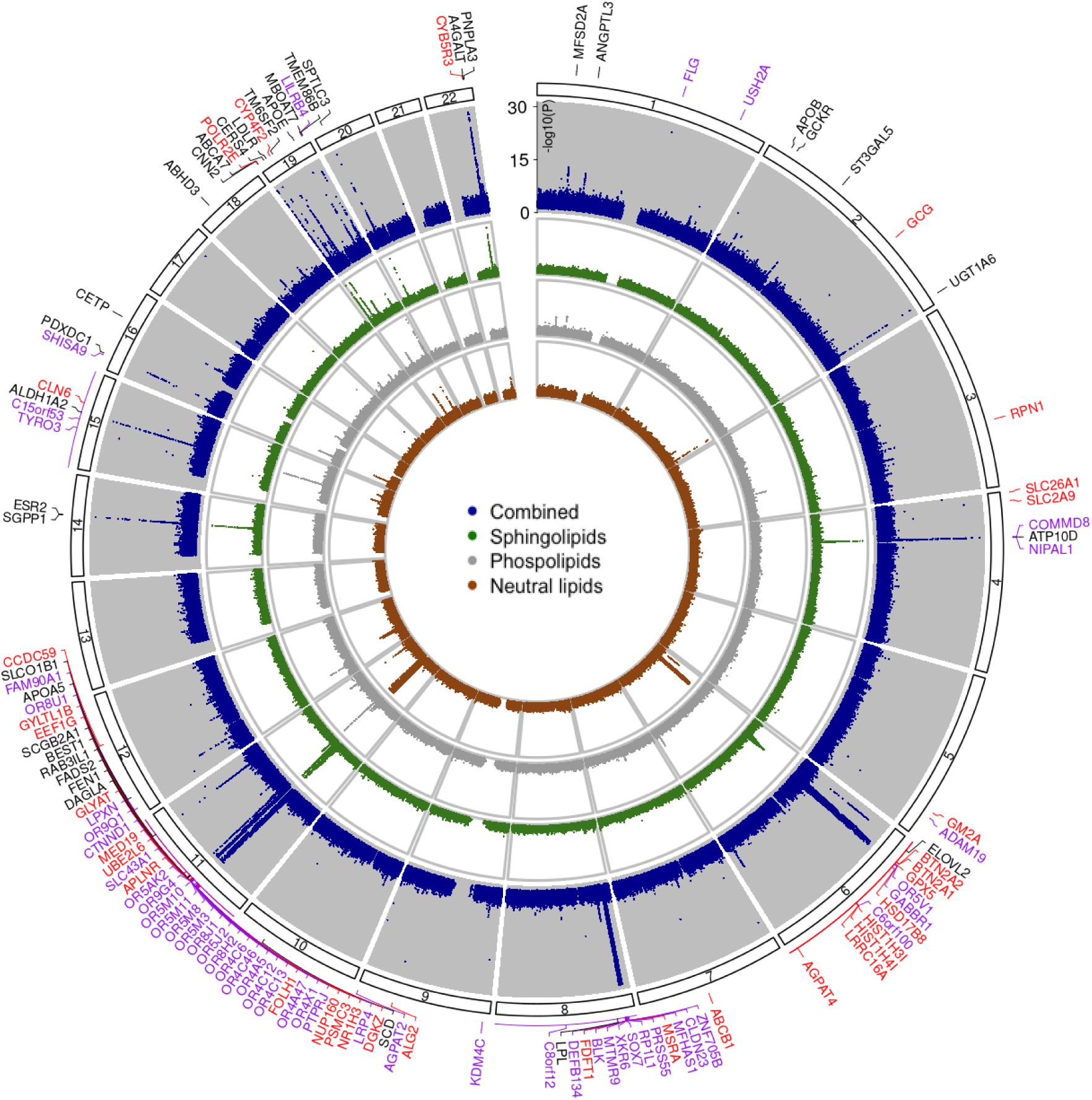
Genomic loci associated with lipid species. Circular Manhattan plot showing the effect loci (adjusted for age, sex and the first 10 genetic principal components), which are coloured black when the lead SNP was identified previously, purple if the lead SNP is novel and linked to the genomic locus based on positional mapping, and red if it is novel and linked to the locus based on in silico functional mapping. Lipid species are subdivided into their main categories (i.e., neutral lipids, phospholipids, sphingolipids, or combined), which are represented by the different circles. The p-value axis is truncated at -log10(p) < 1e-30 for visualization purposes.

### GWAS of fatty acid composition

To evaluate the genetic determinants underlying biochemical features of lipids, we next considered the FA composite measures (n = 267). FA composite measures summarize the concentrations of all lipid species within a class of a certain length and degree of saturation (e.g., DAG(FA18:3)) (**Supplementary Table 3**). Additionally, we utilized information on the total FA composition. These total FA composites included all lipid species, regardless of class, of a specific length and degree of saturation (e.g., FA14:0).

In the GWAS on FA composite measures, we identified 51 genomic loci, of which 25 were novel, after adjusting for sex, age and the first 10 genetic principal components. Some of these were specific to a certain FA tail length or degree of saturation, while others were associated with lipids in general (**Figure 2**). For example, the *FADS2, FADS3* and *ZNF259* (chromosome 11) and *APOE* (chromosome 19) loci were associated with FA composites of almost all lengths and degrees of saturation. In contrast, other loci were associated with lipids of a specific FA length, such as *PKD2L1* on chromosome 10 (16 carbons), *SGPP1* on chromosome 14 (14 and 22 carbons), *PDXDC1* on chromosome 16 (20 carbons), and *A4GALT* on chromosome 22 (22 and 24 carbons) (**Supplementary Table 4**). Similarly, some loci were unique to saturated FAs (e.g., *OR4C12*, chromosome 11), monounsaturated FAs (e.g., *C15ORF53*, chromosome 15; *CPS1*, chromosome 2; *PKD2L1*, chromosome 10), or both saturated and monounsaturated FAs (e.g., *ABCA7*, chromosome 19), whereas others were unique to polyunsaturated FAs, such as *SUGP1* (chromosome 19) (**Supplementary Table 5**). Across all different lengths, lipids carrying a FA tail of 24 (81.2%), 20 (74.2%) or 22 (63.6%) carbons had the highest percentage of significant genetic associations (**Figure 2**).

**Figure 2:**
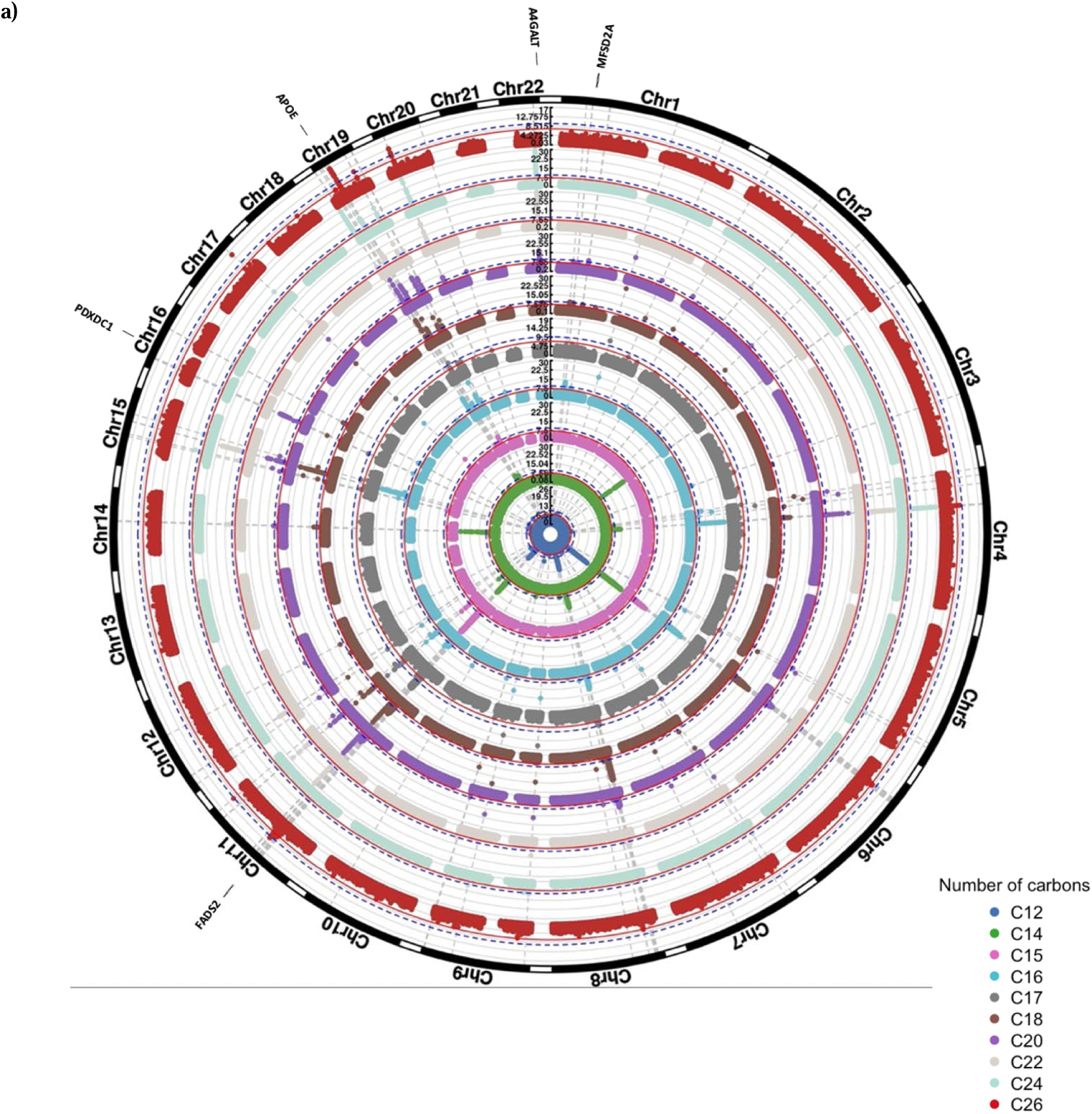
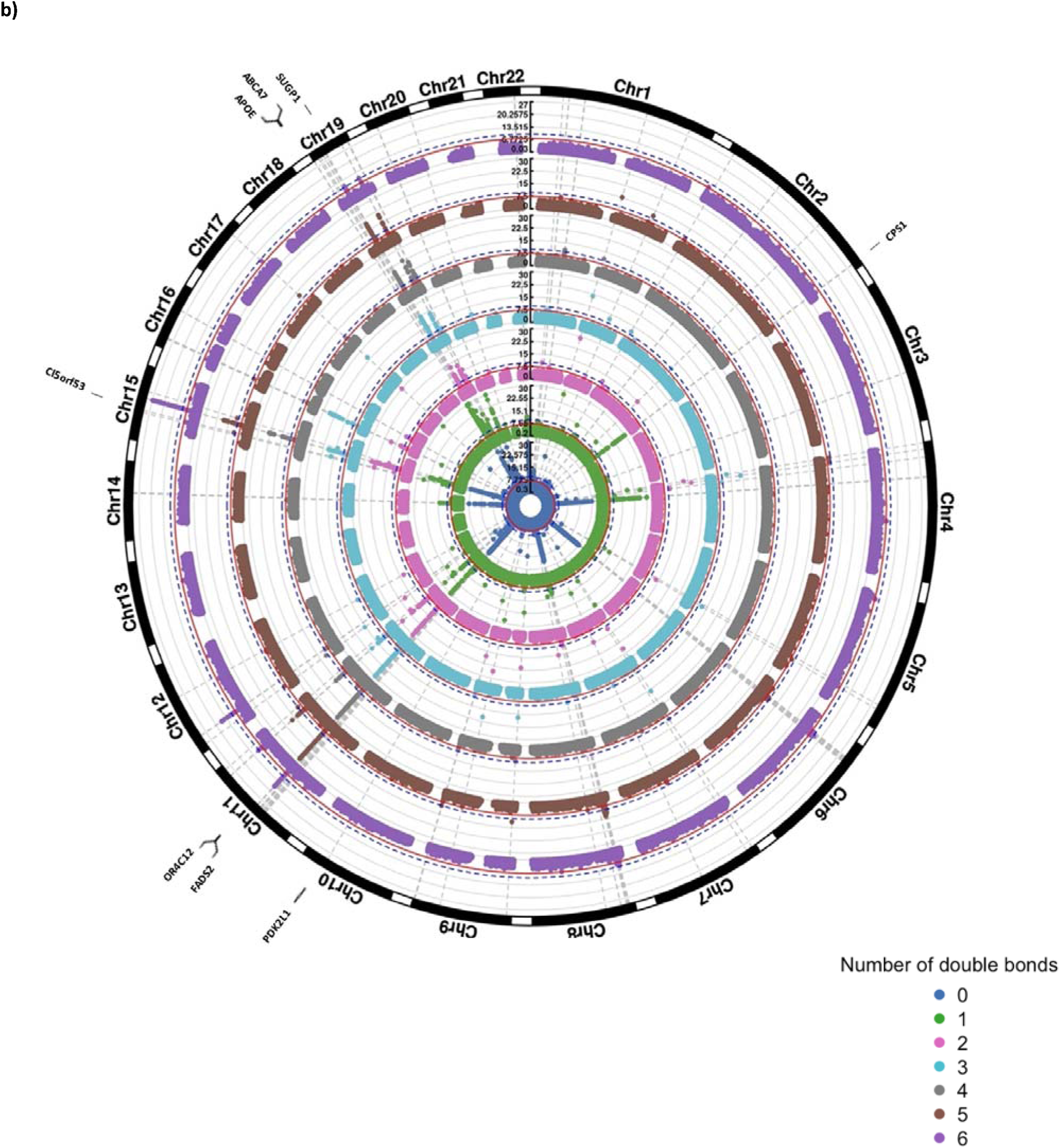
Genetic associations of fatty acid composition. **(a)** Results of the GWAS on fatty acid composition based on the number of carbons. The Manhattan plots are shown for the different fatty acid tail lengths separately, where the inner and outer circles represent the results for all lipids carrying fatty acid tails of 12 or 26 carbons, respectively. **(b)** Results of the GWAS on fatty acid composition based on the number of double bonds. The Manhattan plots are shown for the different fatty acid tail saturations separately, where the inner circle represents the results for all lipids carrying a saturated fatty acid tail and the outer circle for all lipids carrying fatty acid tails with 6 double bonds. All results were adjusted for age, sex and the first 10 genetic principal components. The p-value axis is truncated at -log10(p) < 1e-30 for visualization purposes.

The genomic loci and their independent significant SNPs identified in the GWAS on FA composite measures can be found in the **Supplementary Data 2**.

### Effect of adjustment for clinical lipid measurements and lipid-lowering medications

All results described above were obtained after adjustment for sex, age, and the first 10 genetic principal components (model 1). To evaluate whether the hits were independent of clinical lipid measurements, and allow direct comparison of our results with those from a prior GWAS on lipids (23), we next additionally adjusted our analyses for HDL-C, total serum triglycerides, and total cholesterol levels, and use of lipid-lowering medication (model 2). After adjustment for clinical lipid measurements and the use of lipid-lowering medication, the effect estimates for the lipid species levels changed in a class-dependent manner (**Figure 3a**). Many SNPs were no longer associated with TAGs, DAGs and PEs, while the effect size estimates for the remaining associations became smaller. This is likely due to the genetic correlation between these classes and the clinical lipids. As shown in **Figure 3b**, there is a substantial overlap between the genetics underlying lipid species belonging to these classes and the different clinical lipid measurements. After additional adjustment for clinical lipid measurements, we identified a total of 61 lipid species-related genomic loci, comprising 1251 metabolome-wide significant SNPs and 217 lead SNPs. Of these 61, 48 were already found in model 1. Newly identified genomic loci from model 2 include *F3, CERS6, PAQR9, SLCO5A1, TTC39B, OR4C16, M6PR, SOAT2, KITLG, HNF1A, ABHD2* and *TM4SF5*.

**Figure 3:**
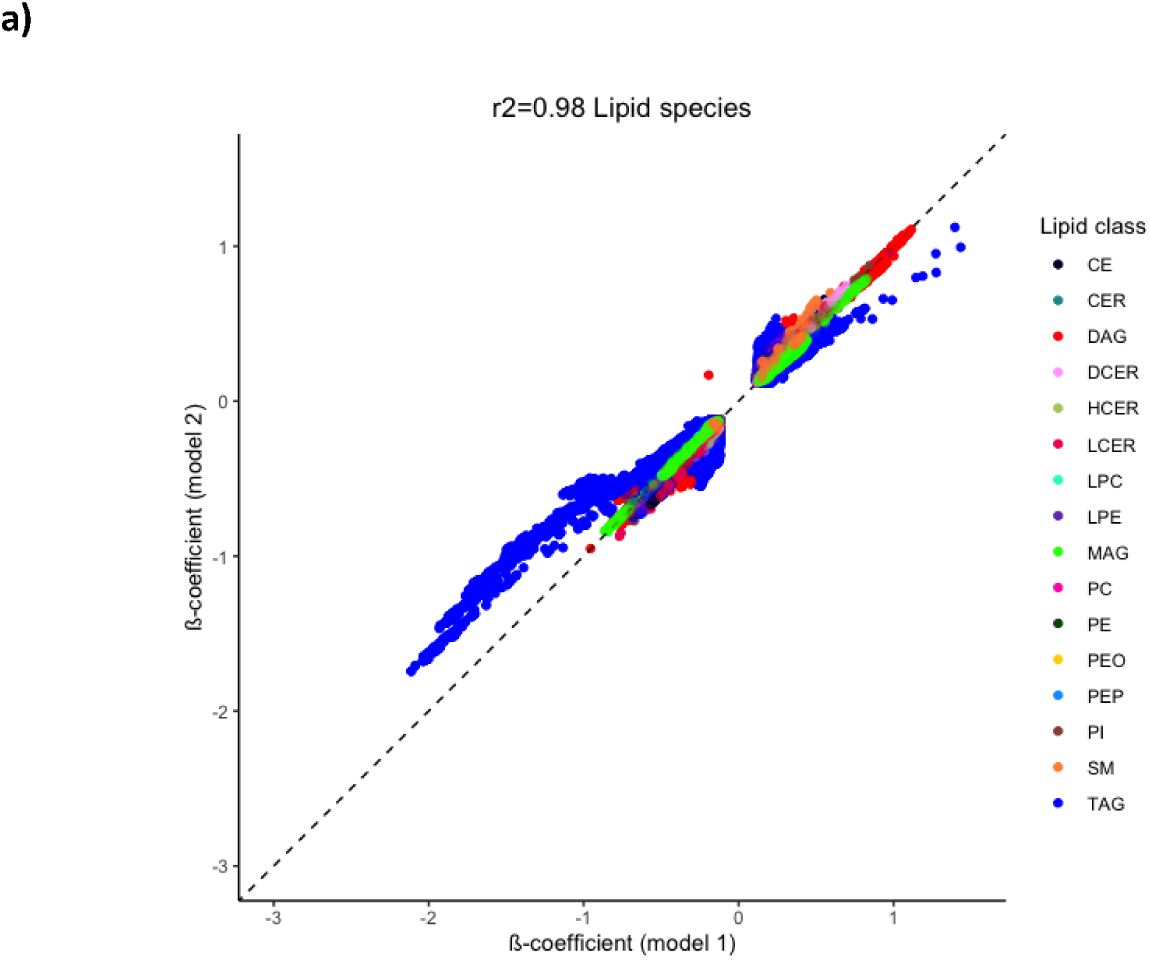
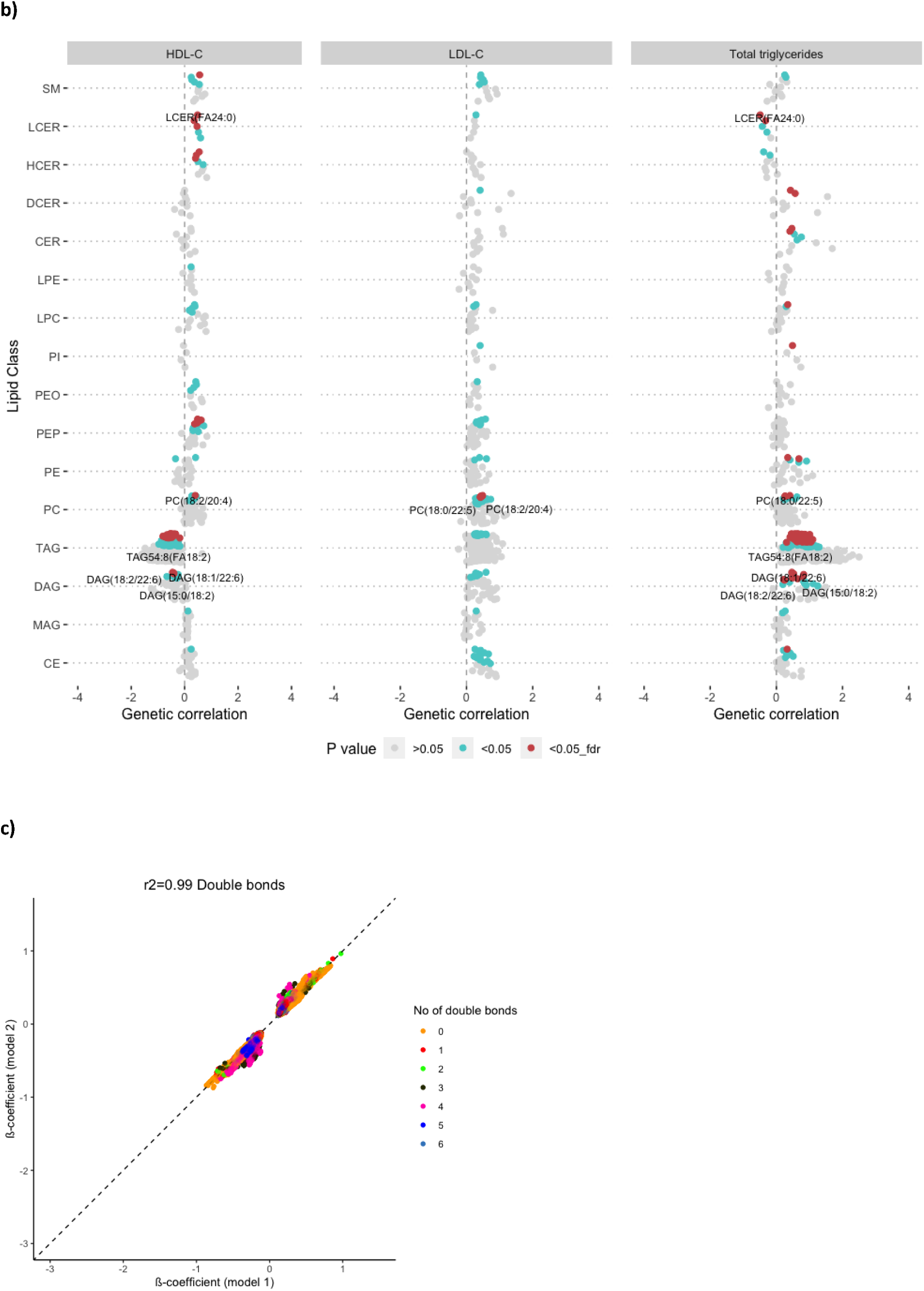
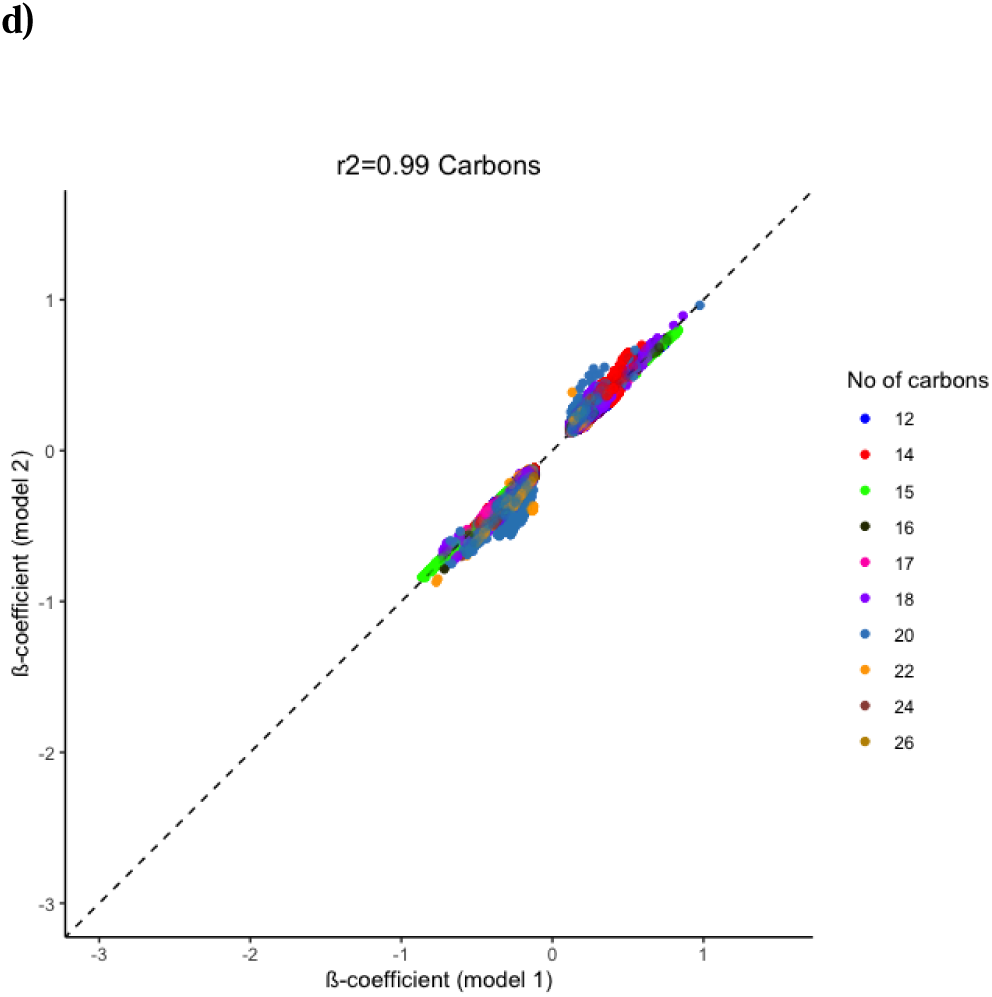
Beta-beta plots and genetic correlations. **(a)** Beta-beta plot depicting the ß coefficients of model 1 (x-axis) against those of model 2 (y-axis) for lipid species. **(b)** The genetic correlation of the lipid species with HDL-C, LDL-C and total triglycerides for model 1. Statistical significance is indicated by colors. The top twenty lipid species based on genetic correlation within each class, which overlapped with at least two clinical lipid measurements, are annotated. **(c)** ß coefficients for model 1 and 2 for lipid composite measures, coloured by the number of carbons, or **(d)** by the number of double bonds. *Abbreviations: HDL-C = high-density lipoprotein cholesterol, LDL-C = low-density lipoprotein cholesterol.*

In contrast, for FA composite measures, the effect estimates across the different biochemical features (i.e., number of carbons and double bonds) remained similar to model 1 (**Figure 3c-d**). There were no changes with regard to the number of carbons or double bonds when comparing the results of model 1 and 2, which aligns well with the small number of significant genetic correlations between these biochemical characteristics and clinical lipid traits (**Supplementary Figure 1**).

All results for model 2 can be found in **Supplementary Data 3** and **4**, followed by model comparisons in **Supplementary Data 5** and genetic correlation results in **Supplementary Data 6**.

### SNP heritability

In order to evaluate how much of the variation in lipid species levels are explained by genetics, we estimated SNP heritability. After adjustment for age, sex, and the first 10 genetic principal components, lipid species showed a median SNP heritability estimate of 0.09 (interquartile range: 0.02 to 0.14) and exhibited substantial inter- and intra-class variability. Generally, we observed that HCERs (median: 0.15, interquartile range: 0.06 to 0.18) and SMs (median: 0.13, inter quartile range: 0.12 to 0.14) had the highest estimated SNP heritability. However, the most highly heritable species did not belong to either of these classes, but to TAGs (TAG36:0(FA12:0), SNP heritability (*h*^2^) =1, SE=0.05) and PEs (PE17:0/18:2, *h*^2^=1, SE=0.66). The least heritable classes were phosphatidylinositols (PIs) (median: 0.00, inter quartile range: 0.00 to 0.01) and lysophosphatidylethanolamines (LPEs) (median: 0.03, inter quartile range: 0.00 to 0.10), but specific species belonging to other classes also showed a low heritability. After adjustment for clinical lipid measurements and use of lipid-lowering medication (model 2), heritability estimates generally decreased (median: 0.04, inter quartile range: 0.00 to 0.10), with MAGs (median: 0.12, inter quartile range: 0.07 to 0.33) and DAGs (median: 0.10, range: 0.03 to 0.12) on average becoming the most heritable, and phosphatidylinositols (PIs) (median: 0.0, inter quartile range: 0.00 to 0. 03) and PEOs (median: 0.02, inter quartile range: 0.00 to 0.10) on average becoming the least heritable **(Supplementary Figure 2a)**. However, there was still substantial variability in SNP heritability of different lipid species across classes.

Next, we estimated the heritability of the FA composite measures separately based on their chain length and degree of saturation. After adjustment for sex, age, and the first 10 genetic principal components, we observed that within MAGs the heritability was higher for saturated FAs (median: 0.63, range: 0.05 to 0.81), compared to unsaturated FAs, where those with six double bonds showed a median heritability of 0.18. On the other hand, cholesteryl esters (CEs) (median heritability: 0.10, range:0.00 to 0.26) carrying a polyunsaturated FA had a higher heritability (median: 0.24, range: 0.09 to 0.24), compared to CEs carrying a saturated (median: 0.08, range: 0.03 to 0.16) or monounsaturated FA (median: 0.05, range: 0.00 to 0.15). Similar to the CEs, TAGs carrying the most polyunsaturated FAs were most heritable (median: 0.16, range:0.13 to 0.18) (**Supplementary Figure 2b**). After additional adjustment for clinical lipid measurements and the use of lipid-lowering medication, we observed that the heritability estimates reduced for the majority of the FA composites, specifically for the CEs (median: 0.02, range: 0.00 to 0.17) and TAGs (median: 0.02, range: 0.00 to 0.16) (**Supplementary Figure 2c**) (17). Nonetheless, a large degree of variability remained across all chain lengths and number of double bonds (**Supplementary Data 7**).

### Annotation of lead SNPs

To disentangle the mechanisms involved in lipid metabolism, we linked the 218 and 217 lead variants from model 1 and model 2 to nearby genes (bottom-up approach – positional) and the ones that had previously been known to be related to metabolic pathways (top-down approach – Gene Ontology (GO), Kyoto Encyclopedia of Genes and Genomes (KEGG), mouse genome informatics (MGI), Orphanet, Reactome pathway databases). Specifically, after adjustment for sex, age, and the first 10 principal components, 497 genes were identified through either the bottom-up or top-down approach. A total of 78 candidate genes overlapped between the two approaches, of which 31 were annotated to novel lead SNPs, and could therefore be classified as candidate genes in lipid metabolism. After additional adjustment for clinical lipid measurements and use of lipid-lowering medication, we identified a total of 506 genes, of which 84 were overlapping (**Supplementary Data 8**). Of these, 28 were annotated to a novel lead SNP. For FA composite measures, we found 64 and 66 candidate genes, of which 46 and 41, respectively, were novel (**Supplementary Data 9)**. Overall, 61 of the identified candidate genes were shared among species and composite measures, which increased to 62 after adjustment for clinical lipid measurements and use of lipid-lowering medication.

We investigated further whether the lead variants were also reported to be associated with gene expression levels in the Genotype-Tissue-Expression (GTEx) resource, and hence could be classified as expression quantitative trait loci (eQTLs). Out of 218 lead variants associated with lipid species levels after adjusting for sex, age, and the first 10 genetic principal components (model 1), 53 were eQTLs, while out of 217 lead SNPs found after adjustment for clinical lipid measurements and use of lipid-lowering medication (model 2), 59 were classified as eQTLs, which largely overlapped with those from the first model. Interestingly, our top hit (rs174547, p-value = 1e-226, beta estimate (β) = −0.626), which was most strongly associated with phosphatidylcholines (PC) (18:0/20:4), was found to be an eQTL for the *FADS2* gene. *FADS2* is a key player in lipid metabolism, regulating fatty acid desaturation, and was also found in several previous GWASs on lipids (1). Our top novel hit (rs1589680, p-value = 1.9e-203, β = −0.637), which was most strongly associated with TAG(36:0(FA12:0), was also an eQTL for *TRIM48*. Additionally, we identified 64 and 82 mQTLs in model 1 and 2, respectively, which were also detected in previous lipidomic GWAS studies.

For the composite measures, 34 eQTLs and 52 mQTLs were identified after adjustment for sex, age, and the first 10 genetic principal components, which changed to 46 eQTLs and 70 mQTLs upon further adjustment for traditional lipid measurements and use of lipid lowering medication. Similar to the results for the lipid species, the top hit (rs174546, p-value = 1.7e-198, β = −0.585) was most strongly associated with PC(FA20:4) and was an eQTL for *FADS2*. Some novel hits were also identified as eQTLs, including rs3747193 SNP (p-value =9.5e-63, β = −0. 492), which was mapped to the *A4GALT* gene and most strongly associated with LCER(FA22:0). The detailed overlap between the associations which were previously reported in metabolomic-based GWAS studies and our lead SNPs are provided in **Supplementary Data 8** and **9**.

### Validation in two independent cohorts

To validate our results, we first used data from a recently published GWAS on lipids in the FinnGen study (n = 7,266) (24). Out of 179 lipid species used in the GWAS from FinnGen, 78 lipid species were also quantified in the Rhineland Study (**Supplemtary Data 12**). Of these, 61 lipid species were associated with at least one SNP at the metabolome-wide significant threshold in the Rhineland Study. For these lipid species, 677 SNP-lipid associations (involving 55 lipid species and 106 SNPs) were nominally significant in FinnGen, while only 76 associations were not. A total of 8,652 associations (involving 1278 independent SNPs and 606 lipid species) could not be tested in FinnGen due to missing data on these SNP-lipid associations in FinnGen (**Supplementary Figure 3a**, **Supplementary Data 1**). Vice versa, data on all 75 lipid species and 111 of FinnGen’s lead SNPs were present in the Rhineland Study. Of the corresponding associations, 175 were replicated in the Rhineland Study, while only 33 were not (**Supplementary Figure 3a**, **Supplementary Data 11**).

We also performed a replication analysis in another independent cohort that used the same lipid panel, namely the EPIC-Potsdam study (n = 1,188) (25). Out of 940 lipid species available in the EPIC-Potsdam study, 915 were also quantified in the Rhineland Study (**Supplementary Data 12**). Out of the 638 and 456 lipid species with at least one independent metabolome-wide significant association in the Rhineland Study in model 1 and model 2, respectively, EPIC-Potsdam had information on 638 and 453. Out of 8,690 associations in model 1, 3,406 were replicated across 565 lipid species and 399 SNPs. We were not able to validate 715 associations across 165 lipid species due to missing data on 135 SNPs in EPIC-Potsdam (**Supplementary Figure 3b, Supplementary Data 1**). In model 2, 2,937 out of 7,209 associations were replicated across 361 lipid species and 325 SNPs. A total of 561 associations spanning 164 SNPs and 178 lipid species could not be validated due to missing data (**Supplementary Figure 3, Supplementary Data 3**). Across all lipid classes, we replicated an average of 73.2% of the SNP-lipid species associations, where the number of SNP-lipid species associations that were replicated within a class ranged from 50% for PEOs to 94.6% for TAGs after adjustment for sex, age, and the first 10 genetic principal components, and from 50% for CER to 100% for PEPs after additional adjustment for clinical lipid measurements and use of lipid lowering medication (**Supplementary Table 6**). Overall, there was a high concordance between results, despite the smaller sample size of the EPIC-Potsdam Study.

### Downstream analysis

The following down-stream analyses are based on the results of the Rhineland Study GWAS conducted on lipid species after adjustment for traditional lipid measures and use of lipid lowering medication (model 2) in order to identify biological pathways underlying the effects of the lead SNPs and the corresponding genes that are independent of known clinical lipid traits.

### Phenome-wide association studies

To further our understanding of how these candidate genes might affect health through lipids, we linked our results to disease phenotypes and clinical measures using PheWASs. We identified several important clusters of variant-trait associations for the 13 lead SNPs for which data was available (**Figure 4**). Firstly, the lead SNP rs1047891 (*CPS1*) was associated with a variety of predominantly cardiometabolic traits and diseases, such as HDL-C levels, systolic blood pressure, glomerular filtration rate, diabetes and chronic kidney failure. Secondly, another important hub centered around rs603424 (*PKD2L*). This lead SNP was linked to phospholipid measurements, blood pressure, chronic kidney disease and cardiovascular diseases among others. Thirdly, rs738409 (*PNPLA3*) was previously linked to several traits, such as serum alanine aminotransferase levels, alcoholic liver cirrhosis, fatty liver disease and precursor cell lymphoblastic leukemia. Finally, although it was not identified as a major hub, the lead SNP rs4147929 (*ABCA7*), which was among the top hits related to fatty acid composition, was associated with Alzheimer’s disease risk. Thus, these genes might affect disease phenotypes through changes in the lipid metabolism. Additional information can be found in **Supplementary Data 10**.

**Figure 4:**
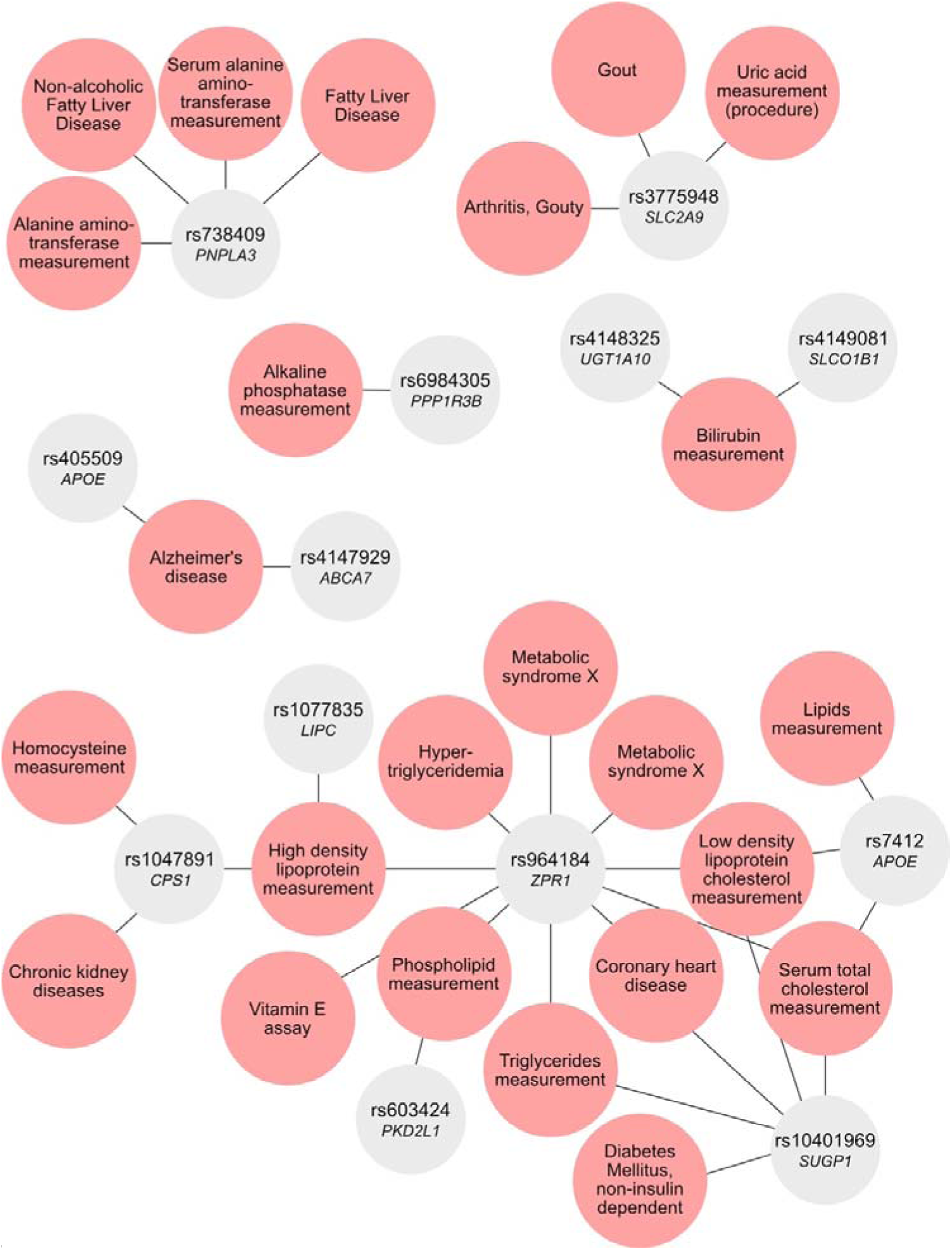
Phenome-wide association studies. The network graph depicts the results of phenome-wide association studies based on the lead single nucleotide polymorphisms (SNPs) available in the DisGeNET database of gene-disease associations, showing phenotype-SNP associations with variant-disease association score > 0.8 and its mapped gene.

### Two-sample Mendelian Randomization analysis of the association between gene expression levels and lipid species

Out of the 506 genes identified through the top-down and bottom-up approaches, whole blood expression quantitative trait loci (eQTLs) were available for 220 genes in GTEx. Overall, through two-sample MR we identified 69 genes that were causally associated with 36 lipid species. We found that the strongest relation was between the expression levels of *TMEM258* and PC(18:0/20:4) (p-value =1.03e-74, β = −2.38, F-statistic= 48.32). Among the novel candidate genes, we found evidence for a causal association of *FDFT1* (p-value =3.69e-24, β =-0.90, F-statistic= 53.85) and *BLK (*p-value =1.25e-27, β =0.98, F-statistic=133.46) expression with DAG(12:0/18:0). Furthermore, the MR analysis indicated a causal association between the expression levels of *MFHAS1* and DAG(12:0/18:0) (p-value =5.34e-8, β =-1.03, F-statistic=27.33) (**Table 2**, **Supplementary Data 13**).

**Table 2:**
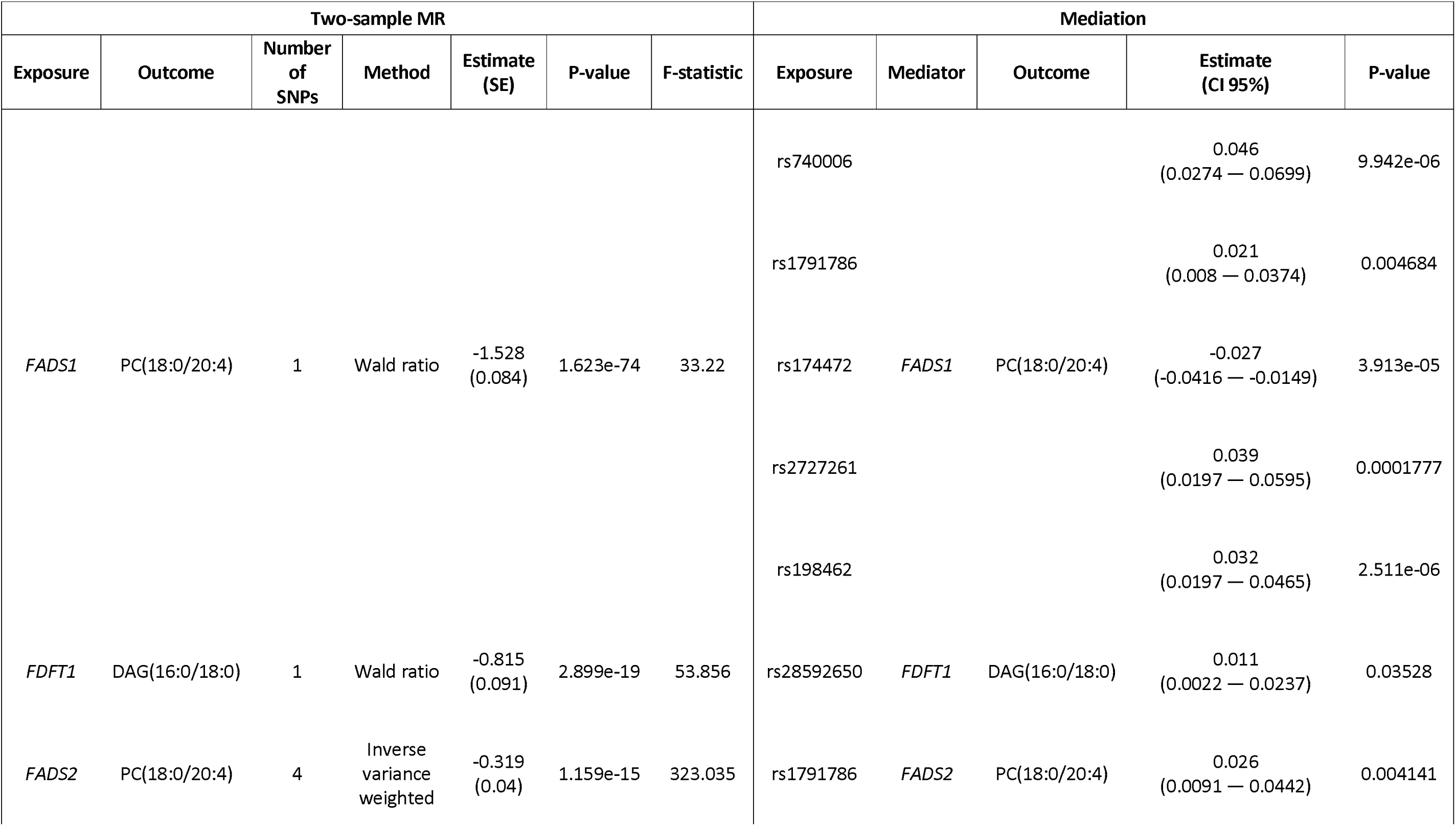
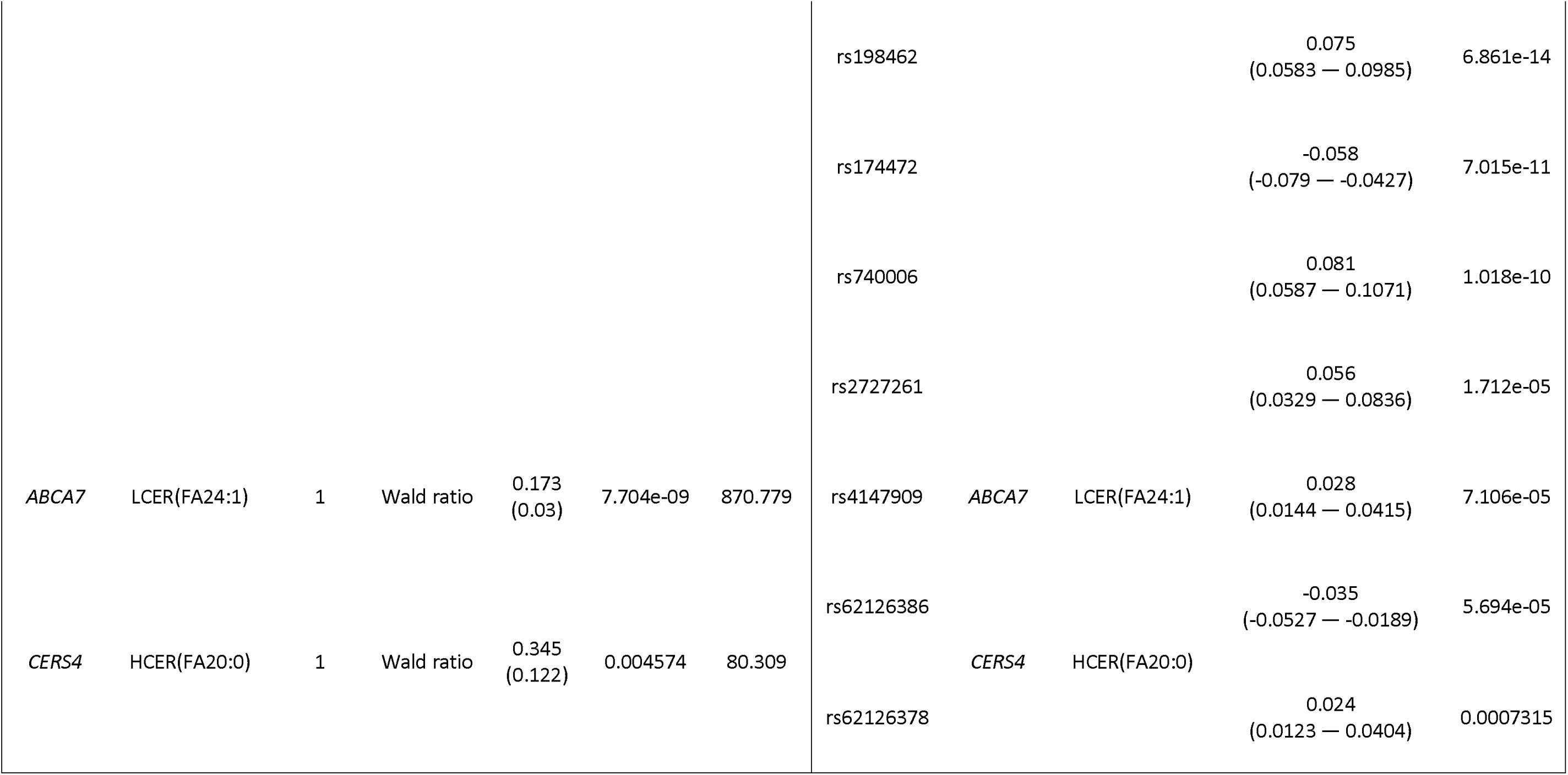
Causal associations between genes and lipid species identified using two-sample Mendelian Randomization (MR), and confirmed through mediation analysis.

### Functional validation through gene expression analysis

We performed functional validation analyses using data from 2,146 Rhineland Study participants for whom transcriptomic and lipidomic data were available. Out of the 506 genes identified through the top-down and bottom-up approaches, the expression levels of 229 were available in the Rhineland Study. The expression levels of 42 of these genes were significantly associated with 27 lipid species. The strongest association was observed between *FADS2* expression and PC(18:0/20:4) (β = −0.238, p-value < 0.001). The eQTL analysis showed 75 significant associations (false discovery rate (FDR) < 0.05) between lead SNPs and expression levels of primary candidate genes, with rs174564 as the top hit associated with *FADS2* expression (β = 1.817, FDR < 0.001) (**Supplementary Data 14**). In addition, we performed mediation analyses using the expression levels of the genes (n = 16) that were significantly associated with both lipid species concentrations and their corresponding lead SNPs. This analysis revealed that the expression levels of several candidate genes partially mediated the SNP-lipid species relationships, such as *FADS2*, which was the strongest mediator of the rs198462 - PC(18:0/20:4) association (β = 0.08, p-value < 0.001, percentage of the effect mediated: 39.5%), and *ABCA7*, mediator of the rs4147909 – LCER(FA24:1) association (β = 0.03, p-value < 0.001, percentage of the effect mediated: 11.5%) (**Table 2**). For novel SNP-lipid species associations, we identified *FDFT1* as a partial mediator (mediated effect: 4.7%) of the association between rs28592650 and DAG (16:0/18:0).

The network-based analysis clustered the primary candidate genes into eight modules, consisting of 78 hub-genes, which were arbitrarily assigned color names. The biological significance of the top modules became clear after querying of the KEGG database, which revealed enrichment for pathways such as glycosaminoglycan degradation, glycerophospholipid metabolism, peroxisome pathways, mucin type O-Glycan biosynthesis and glycerolipid metabolism (**Supplementary Data 15**).

## Discussion

To elucidate the genetic architecture of circulating complex lipid levels and composition we conducted a comprehensive GWAS encompassing a wide variety of lipid species and plasma FA composite measures. In the Rhineland Study (n= 6,096), we identified a total of 57 genomic loci before adjustment for routine clinical lipid measurements (i.e., total serum triglycerides, HDL-C, and total cholesterol levels) and medication, and 61 after additional adjustment for these measures. Importantly, around half of the identified genomic loci were novel. Moreover, we discovered 84 candidate genes that were associated with lipid species independent of routine clinical lipid measurements and medication. By comparing our results to a previously published GWAS on a smaller set of lipids (FinnGen, n = 7,266), we found that we could replicate almost all of the previously reported associations (84.1%). Conversely, almost all of our metabolome-wide significant results, for which data were available in FinnGen, were replicated in this independent cohort (89.9%). We further demonstrated the robustness of our findings by running a second replication analysis in an independent cohort with the same lipid panel (EPIC-Potsdam, n = 1,188). Despite the smaller sample size, around 39.2% of our associations were replicated in the EPIC-Potsdam cohort as well.

Importantly, we did not only assess lipid species levels, but also the overall FA composition of the lipidome, demonstrating that some genes were specific to a certain biochemical feature (i.e., FA tail length or degree of saturation), such as *SGPP1* and *A4GALT*, while others were associated with a variety of composite measures across the entire lipid spectrum, such as *FADS2* and *APOE*. We also found that heritability of complex lipids showed high variability within classes and was mostly determined by a lipid’s tail length as opposed to its class or degree of saturation. Although lipid species within the sphingolipids and neutral lipids showed substantial genetic overlap with clinical lipids, in line with previous studies (17), the genetic overlap with the number of carbons and double bonds was considerably lower. This lack of genetic overlap could account for our finding that the effect sizes of SNPs associated with levels of lipids belonging to these categories became non-significant upon adjustment for clinical lipids, while the effects of genetic variants associated with biochemical features of lipids were robust to further adjustment. Interestingly, our PheWAS analysis revealed links between the genetic modifiers of complex lipids and several diseases, including cardiovascular diseases and Alzheimer’s disease, as well as other metabolic traits, including levels of HDL-C and phospholipids. Next, our two-sample MR analyses yielded evidence for causal associations between the expression levels of a number of candidate genes identified in our *in silico* analysis and complex lipid levels. Moreover, our functional analysis demonstrated that up to 36.7% of the effects of several lead SNPs on lipid species were mediated through changes in gene expression. Finally, the pathway analysis provided further corroboration of our results by demonstrating that many of the mapped genes were indeed involved in pathways related to lipid metabolism.

Importantly, in contrast to most previous genetic association studies of complex lipids, we evaluated the genetic architecture of the whole plasma FA composition as well as the shared biochemical properties across different lipid species and classes, including their tail length and degree of saturation. By assessing FA composite measures, we could identify specific genes uniquely associated with certain biochemical properties of lipid composites. While genes like *FADS2*, *APOE* and *FLG* were associated with many complex lipids regardless of their precise biochemical properties, we found that others, such as *ABCA7*, were predominantly related to lipids with a specific degree of saturation such as with LCERs that were either saturated or monounsaturated FAs. Similarly, *PDXDC1* was related to lipids with specific tail lengths, such as with FAs carrying 20 carbons in LPCs, CEs and LPEs classes. In addition, we found that the heritability of FA composition was mainly determined by class or degree of saturation rather than by the lipid’s tail length. This was mainly observed for MAGs, where heritability was higher for saturated FAs and lower for unsaturated FAs, whereas for CEs and TAGs we observed the opposite. Collectively, our findings thus indicate that genetics play a crucial role in determining both the levels and biochemical attributes of complex lipids, extending previous findings that both aspects are pivotal for the effects of different lipid species (7,26–28).

Using two-sample MR, we identified 69 genes that were potentially causally associated with 36 lipid species. Among them, we demonstrated that the expression levels of *FADS2*, *FADS1*, *CERS4*, *ABCA7* and *FDFT1* also partially mediate several associations between lead SNPs and lipid species’ levels. Both *FADS2* and *FADS1* have a critical role in FA metabolism, which is further supported by our results (29,30). *CERS4* was most consistently associated with total concentrations of sphingolipids, as well as individual sphingolipid species’ levels and various FA lengths, which is biologically highly plausible given that *CERS4* encodes a ceramide synthase. Moreover, one previous study also specifically reported an association of genetic variants in *CERS4* with FA20:0, which we confirmed and expanded upon by showing an association with FA18:0 as well (31).

Interestingly, *ABCA7* was one of the most significant hits related to FA composite measures, especially those involving monounsaturated and saturated FAs, where its expression levels partially mediated the SNP-lipid associations. *ABCA7* encodes a lipid transporter and has previously been associated with an increased risk of Alzheimer’s disease (32,33). Moreover, recently a potential causal role of LCERs in the relation between *ABCA7* and the risk of Alzheimer’s disease was reported (34). Our MR and mediation results extend these findings, indicating a pivotal role for *ABCA7* in the regulation of lipid metabolism, where its expression particularly controls monounsaturated and saturated lipids. Thus, we not only discovered a large number of mQTLs of complex lipids, but also pinpoint genes whose expression levels are likely to causally mediate these associations.

Results of our PheWAS further highlight the important role of complex lipids as modifiers of a large number of different traits and diseases. For example, genetic variants in *CPS1* have previously been associated with a variety of different phenotypes, including HDL-C levels and diabetes (35). We found that *CPS1* was specific to monounsaturated cholesteryl esters, which are intimately linked to HDL-C as well as diabetes and other diseases (13,36). Another hub gene, *PKD2L*, has previously been associated with cardiometabolic health as well as phospholipid measurements among others. We extend these findings by showing that *PKD2L* mainly affects monounsaturated lipids. *PNPLA3*, on the other hand, was mainly associated with MAGs and TAGs in our analysis. Interestingly, this gene was previously reported in relation to alcoholic liver cirrhosis and fatty liver disease, which are characterized by an increase in triglycerides in the liver (37). Moreover, some of our hits have also been linked to aspects of lipid metabolism before, such as the “biosynthesis of unsaturated FAs”, which further strengthens our results. Thus, the PheWAS findings demonstrate that the genes related to our lead SNPs are at the center of different cardiometabolic traits and diseases and are involved in the regulation of specific aspects of lipid metabolism. Therefore, they might serve as potential therapeutic targets for restoring disturbed lipid metabolism.

Our study has both strengths and limitations. First, although we present one of the largest and most comprehensive GWAS studies on complex lipid levels and composition to date, we did not perform experimental validation of our findings in model systems. However, our systematic functional genomics approach identified several candidate genes, a large number of which we could functionally validate by using matched individual-level transcriptomics data. Moreover, we performed rigorous validation of our results leveraging data from two other independent cohorts. The extensive unique complex lipids data on a large number of individuals is a main strength of our study, as we could investigate the genetics of specific species, as well as the whole FA composition, which had been challenging so far. Nonetheless, despite the platform’s large coverage, some lipids and lipid classes were not included, such as cardiolipins, phosphatidylserines and gangliosides. Furthermore, the complete biochemistry of certain lipids could not be confirmed, such as the total composition of the TAGs or the stereochemistry of FA tails.

In conclusion, we unraveled the genetic basis of a wide variety of circulating lipid species as well as FA composition. We discovered a large number of novel mQTLs for complex lipids, identified candidate genes that could be crucial in regulating specific aspects of lipid metabolism, and provide evidence for their clinical relevance by linking them to a diverse array of cardiometabolic traits and diseases.

## Methods

### Study population

We used data from the Rhineland Study, an ongoing prospective cohort study in Bonn, Germany. People aged 30 or above who lived in either of two geographically defined areas in Bonn were invited to participate. Participants are invited in random order, based on postal code. The only exclusion criterium was having insufficient command of the German language to provide informed consent. Participants underwent deep phenotyping to obtain genetic, imaging, socio-demographic, clinical as well as metabolic information. Participants who enrolled before the 26^th^ of November 2021 (n= 8,318) were included when they had complete lipidomic and genetic data (n= 6,096).

In addition, data from the independent EPIC-Potsdam study was used for a replication analysis. EPIC-Potsdam is a prospective cohort study in Potsdam, Germany, that started in 1994 and includes ∼27,500 participants between the ages of 35 and 64 at baseline (25). Data was collected on diet, anthropometrics, genetic and lipidomic measures, among others. In the current analysis, all participants from a random subsample (n = 1,188, ∼5% of total participants) with complete genetic, lipidomic, and clinical lipid data available were included.

### Lipidomics

Absolute (nmol/ml) plasma lipid concentrations were measured using the Metabolon Complex Lipids platform. An automated Butanol–Methanol (BUME) method was used for lipid extraction (38). First, extracts were dried and reconstituted in ammonium acetate dichloromethane:methanol which was then transferred for analysis in a Shimadzu Liquid Chromatography with nano PEEK tubing and the Sciex Selexlon-5500 QTRAP. To extract various different lipids, both positive and negative mode electrospray were used. By multiplying the concentration of the internal standard with the ratio between the signal intensity of the target compound and the internal standard, the absolute concentration of 1,023 lipid species was quantified, grouped into neutral, phospho- and sphingolipids based on their fundamental biochemical structural properties (39). Besides individual species, FA composite measures (n = 271) were also calculated. FA composite measures summarize the concentrations of all lipid species that have at least one FA tail of a specific length and saturation. Lipid species were characterized by a complete identification of all FA tails for all lipid classes except TAGs (e.g. DAG(16:1/20:0, CER20:1) (**Supplementary Table 4**). For the TAGs, which have 3 FA tails, lipids were considered species when information on the total number of carbons and double bonds as well as detailed information on one FA tail was available (e.g. TAG53:5(FA18:3)). A more detailed overview with examples of the lipid nomenclature used in this paper can be found in **Supplementary Table 4**. Only lipids present in at least 1% of the population were evaluated in the current analysis (n= 970).

### Genomics

Blood samples were genotyped using the Illumina Omni-2.5 exome array, which contains 2,612,357 single-nucleotide polymorphisms (SNPs). Subsequently, genotype data were processed using GenomeStudio (version 2.0.5), and quality controlled with PLINK (version 1.9). SNPs were excluded if not meeting the Hardy-Weinberg disequilibrium criterium (p-value < 1E-5), having a minor allele frequency < 0.01, or showing a poor genotyping rate (< 99%). Additionally, participants were excluded because of a poor call rate of less than 95% (n = 41), abnormal heterozygosity (n = 69), cryptic relatedness (n = 261), or a sex mismatch (n = 28). To account for variation in population structure, which may otherwise cause systematic differences in allele frequencies (40), we used EIGENSTRAT (version 16000) (41). EIGENSTRAT uses principal components to detect and correct for population structure, which resulted in the exclusion of an additional 164 participants from non-central European descent. Finally, we imputed missing SNPs with IMPUTE (version 2) (42) based on the 1000 Genomes (phase 3) reference panel (43). We ensured a high imputation quality by only including SNPs with an info score metric > 0.3, which indicates reliable imputation (44).

### GWAS

We performed a GWAS on the absolute concentrations of lipid species and FA composite measures using Rvtests (42). In the first model, we regressed out the effects of age, sex and the first 10 genetic principal components to account for population structure. In the second model, we additionally adjusted for the levels of total cholesterol, triglycerides, and HDL-C, as well as the use of lipid-lowering medication. Residuals obtained in either model were subsequently normalized using a rank-based inverse normal transformation before performing the GWAS. In the second model, only the participants with complete data on the additional covariates were included. We set the genome-wide significance level at p < 5E-8. Because of the large number of outcome variables (i.e., adjusted metabolite levels), we corrected for multiple testing using the method of Li et al. (44), which accounts for the correlation between the lipid levels by estimating the effective number of independent tests. Accordingly, we estimated the effective number of independent tests to be 222, resulting in a metabolome-wide significance level of p < 2.27E-10 (≈ 5E-8/222).

For further functional mapping of genomic loci that influence lipid metabolism, we aggregated all GWAS results in a unique dataset, in which we included the lowest p-value across all lipids for each SNP as described previously (23). Next, the aggregated data were processed with the Functional Mapping and Annotation (FUMA) tool (45) to identify the lead and independent SNPs associated with lipid species and FA composite measures. We used the 1000 Genomes dataset (phase 3) as a reference panel to account for the linkage disequilibrium (LD) structure of the metabolome-wide significant genetic variants and identification of independent metabolome-wide significant SNPs (squared allelic correlation (r^2^) ≥ 0.6). Independent lead metabolome-wide significant SNPs were defined as those genetic variants with r^2^ ≥ 0.1. Each genomic risk locus was defined by tagging independent significant SNPs in close physical proximity (i.e., within 250 kb from either side of each LD block). The genomic risk loci thus contain multiple independent significant and lead SNPs.

### Annotation of lead SNPs

We systematically identified putative causal genes involved in lipid metabolism using both the top-down and bottom-up approaches within the ProGeM framework (46). The top-down approach uses the curated database of known metabolic-related genes to identify biologically relevant genes that are present within 500 kb of the lead SNP. The bottom-up approach identifies genes based on genomic distance, where the three closest protein-coding genes near the lead SNP are selected. We then used the impact factor score, as calculated by the Variant Effect Predictor (VEP) method (47), to classify the lead SNP’s function as either missense, start loss or stop gain. Primary candidate genes for lipid metabolism were defined as those genes that were identified through both the top-down and the bottom-up approach. We also used mGWAS-Explorer (48) and PhenoScanner (49) to assess whether our lead SNPs had been identified previously in other lipidomic GWAS studies.

### Genetic architecture of complex lipids

To investigate the genetic heritability of the lipid species and FA composition, we calculated the SNP-based heritability using Genome-wide Complex Trait Analysis (GCTA) (50). For this, we took the rank-based inverse-transformed residuals of lipid species and FA composites as phenotypes obtained from model 1 and model 2 in the Rhineland Study. In addition, we used LD Score Regression (51) to calculate the genetic correlation between different complex lipids. For the genetic correlation analysis, we used the summary statistics from the largest GWAS studies on three traditional lipid traits (52–55) (including HDL-C, LDL-C and triglycerides). A false discovery rate (FDR) < 0.05 was considered statistically significant.

### Validation of GWAS results in independent cohorts

First, we performed validation of our GWAS results using summary statistics derived from a recently published large scale GWAS study on complex lipids, including 179 lipid species, in the FinnGen cohort (24). We compared whether our metabolome-wide significant SNPs replicated at a nominally significant level (p < 0.05) in the FinnGen GWAS summary statistics. In addition, we evaluated whether the lead SNP associations reported in the FinnGen cohort replicated in our study. Second, we validated our results by running a replication analysis in the EPIC-Potsdam study, which uses the same lipidomic platform. Similar to the approach above, we checked whether our metabolome-wide significant SNPs were nominally significant (p < 0.05) in a separately-run GWAS on complex lipids in the EPIC-Potsdam cohort.

### Phenome-wide association studies

To further assess the clinical relevance of our findings, we also performed PheWAS analyses. We used the DisGeNET database to assess the relation between the lead SNPs associated with lipid species, after adjustment for traditional lipid measures, and the aforementioned phenotypes. Using the disgenet2r package, we obtained Variant-Disease Association (VDA) scores, which translates the certainty of the results into a score ranging from 0 to 1 (56). We only report the phenotype-SNP associations that had a VDA score > 0.2.

### Two-sample Mendelian Randomization

We employed a two-sample MR approach to test whether the associations between the genes identified in our top-down and bottom-up approaches and lipid species (adjusted for traditional lipids measures) were causal, using the TwoSample MR package. We downloaded whole blood eQTL data from the GTEx resource (57), and used variants associated with gene expression levels as genetic instruments in the MR analyses. Specifically, eQTLs of candidate genes were used as the exposure and variants associated with lipid species levels as the outcome. We clumped SNPs in LD (r^2^ < 0.01 within a 10 Mb window). The method selection for Two-sample MR was based on the number of SNP instruments available, including Wald Ratio (1 SNP instrument), inverse variance weighted method (>=2 SNP instruments), and MR-Egger method (>3 SNP instruments). To assess the robustness of our MR results, we performed several sensitivity analyses, such as calculating F-statistics to assess the risk of weak-instrumental bias and MREgger intercept to identify horizontal pleiotropy.

### Functional validation through gene expression analysis

Lastly, we functionally validated our results using gene expression data of the first 3000 consecutively enrolled participants of the Rhineland Study. Samples used for RNA sequencing were stored in PAXgene Blood RNA tubes (PreAnalytix/Qiagen), and were thawed and incubated at room temperature to increase RNA yields. Total RNA was isolated according to manufactures’ instructions using PAXgene Blood miRNA Kit following their automated purification protocol (PreAnalytix/Qiagen). RNA integrity and quantity were evaluated using the tapestation RNA assay on a Tapestation4200 instrument (Agilent). The tapestation RNA assay (Tapestation4200 instrument from Agilent) was used to assess RNA integrity and quantity. After using 750 ng of total RNA to generate next generation-sequencing libraries for total RNA sequencing (TruSeq stranded total RNA kit,Illumina), a Ribo-Zero Globin reduction was performed. Libraries were quantified using Qubit HS dsDNA assay (Invitrogen) and clustered at 250 pM concentrations on a NovaSeq6000 instrument. Quality control of the sequencing reads was performed with FastQC (v0.11.9). Following the filtering of low-quality score reads with Trimmomatic (v.0.39), sequencing reads were aligned to the human reference genome GRCh38.p13 using STAR (v2.7.1). The count matrix was generated with STAR –quantMode GeneCounts using the human gene annotation version GRCh38.101. Genes with overall mean expression > 15 reads and expressed in at least 5% of the participants were retained for further analysis. Raw counts were normalized and transformed using the varianceStabilizingTransformation function from DESeq2 (v1.30.1).

For functional validation of the primary candidate genes, we first investigated the association of lipid species levels (dependent variable) with the residuals of candidate genes’ expression levels after adjustment for age, sex, batch, red and white blood cell counts, as well as the fraction of basophils, eosinophils, lymphocytes, monocytes, and neutrophils (independent variables) using linear regression. The analysis was conducted on data from 2,146 participants of the Rhineland Study for whom gene expression and lipid species data were available. For this validation step we set the statistical significance threshold at p < 0.05. Secondly, we conducted an eQTL analysis for the lead SNPs with the residuals of candidate genes’ expression levels after further adjustment for the first 10 genetic PCs including participants with complete gene expression and genotype data (n = 2,008), setting the statistical significance level at FDR < 0.05. Lastly, we performed mediation analyses using the R package lavaan (v.0.6-11) to investigate to what extent gene expression levels (mediator) mediated the relation between lead SNPs (exposure) and lipid species levels (outcome).

To identify key modules and hub-genes involved in complex lipid metabolism, we performed a weighted gene correlation network analysis of the primary candidate genes using the R package WGCNA (v.1.70-3) (58). Briefly, gene expression levels were adjusted for batch effects and the residuals were used to construct an adjacency matrix based on signed bi-weighted mid-correlations, using a soft threshold power of 5. Co-expressed genes were then clustered as modules using cutreeDynamic function (method = hybrid, minimum module size = 5, deepSplit = 3, pamRespectsDendro = TRUE). Finally, we merged the modules which had similar profiles using a height cut of 0.2. Eigengenes were calculated for each module and adjusted for age, sex, red and white blood cell counts, and the fraction of basophils, eosinophils, lymphocytes, monocytes, and neutrophils. The enrichment pathways for the different modules were subsequently assessed based on KEGG using the WEB-based Gene SeT AnaLysis Toolkit (59). Genes with a value of “geneModuleMembership” greater than 0.60 were considered hub-genes for the corresponding modules.

## Author Contributions

**Elvire N. Landstra:** Conceptualization, Methodology, Formal Analysis, Writing – Original Draft Preparation, Visualization; **Mohammed A. Imtiaz:** Conceptualization, Methodology, Formal Analysis, Writing – Original Draft Preparation, Visualization; **V. Talevi:** Conceptualization, Methodology, Formal Analysis, Writing – Original Draft Preparation, Visualization; **F. Eichelmann:** Data Curation, Formal Analysis, Writing – Reviewing and Editing; **M.B. Schulze:** Data Curation, Resources, Writing – Reviewing and Editing; **N. Ahmad Aziz:** Conceptualization, Methodology, Writing – Reviewing and Editing, Data Curation, Supervision; **Monique M.B. Breteler:** Conceptualization, Methodology, Resources, Writing – Reviewing and Editing, Data Curation, Funding Acquisition, Supervision.

## Ethical approval

Approval to undertake the Rhineland Study was obtained from the ethics committee of the University of Bonn, Medical Faculty. The study was carried out in accordance with the recommendations of the International Council for Harmonisation Good Clinical Practice standards. We obtained written informed consent from all participants in accordance with the Declaration of Helsinki.

The study protocol of EPIC-Potsdam was approved by the ethics committee of the Medical Society of the State of Brandenburg, Germany, and all participants provided a statement of written informed consent before enrolment.

## Acknowledgements

We would like to thank all participants and the study personnel of the Rhineland Study and EPIC-Potsdam. The Rhineland Study is funded by the German Center for Neurodegenerative Diseases (DZNE). This work was further supported by the Diet-Body-Brain Competence Cluster in Nutrition Research funded by the Federal Ministry of Education and Research (grant number 01EA1410C), the Federal Ministry of Education and Research (FKZ :01KX2230) in the framework “PreBeDem - Mit Prävention und Behandlung gegen Demenz”, the Deutsche Forschungsgemeinschaft (DFG, German Research Foundation) under Germanýs Excellence Strategy – EXC 2151 – 390873048, the Deutsche Forschungsgemeinschaft (DFG, German Research Foundation) – SFB 1454 – Project-ID 432325352, and the Helmholtz Association under the 2023 Innovation Pool. The work in EPIC-Potsdam was supported by a grant from the European Commission and the German Federal Ministry of Education and Research within the Joint Programming Initiative A Healthy Diet for a Healthy Life, within the ERA-HDHL cofounded joint call Biomarkers for Nutrition and Health (01EA1704), grants from the German Federal Ministry of Education and Research and the State of Brandenburg to the German Center for Diabetes Research (DZD) (82DZD00302, 82DZD03D03).

N.A. Aziz is supported by a European Research Council Starting Grant (Number: 101041677).

## Data availability

The data from the Rhineland Study and EPIC-Potsdam used in this manuscript are not publicly available due to data protection regulations. Access to data can be provided to scientists in accordance with the Rhineland Study’s Data Use and Access Policy. Requests for additional information and/or access to the datasets of the Rhineland study can be send to. Information on data access and contact details for the EPIC-Potsdam study can be obtained at https://www.dife.de/en/research/cooperations/epic-study/. All authors had full access to the data and take responsibility for the integrity of the data as well as the accuracy of the analysis.

## Extended tables and figures

**Extended Table 1:**
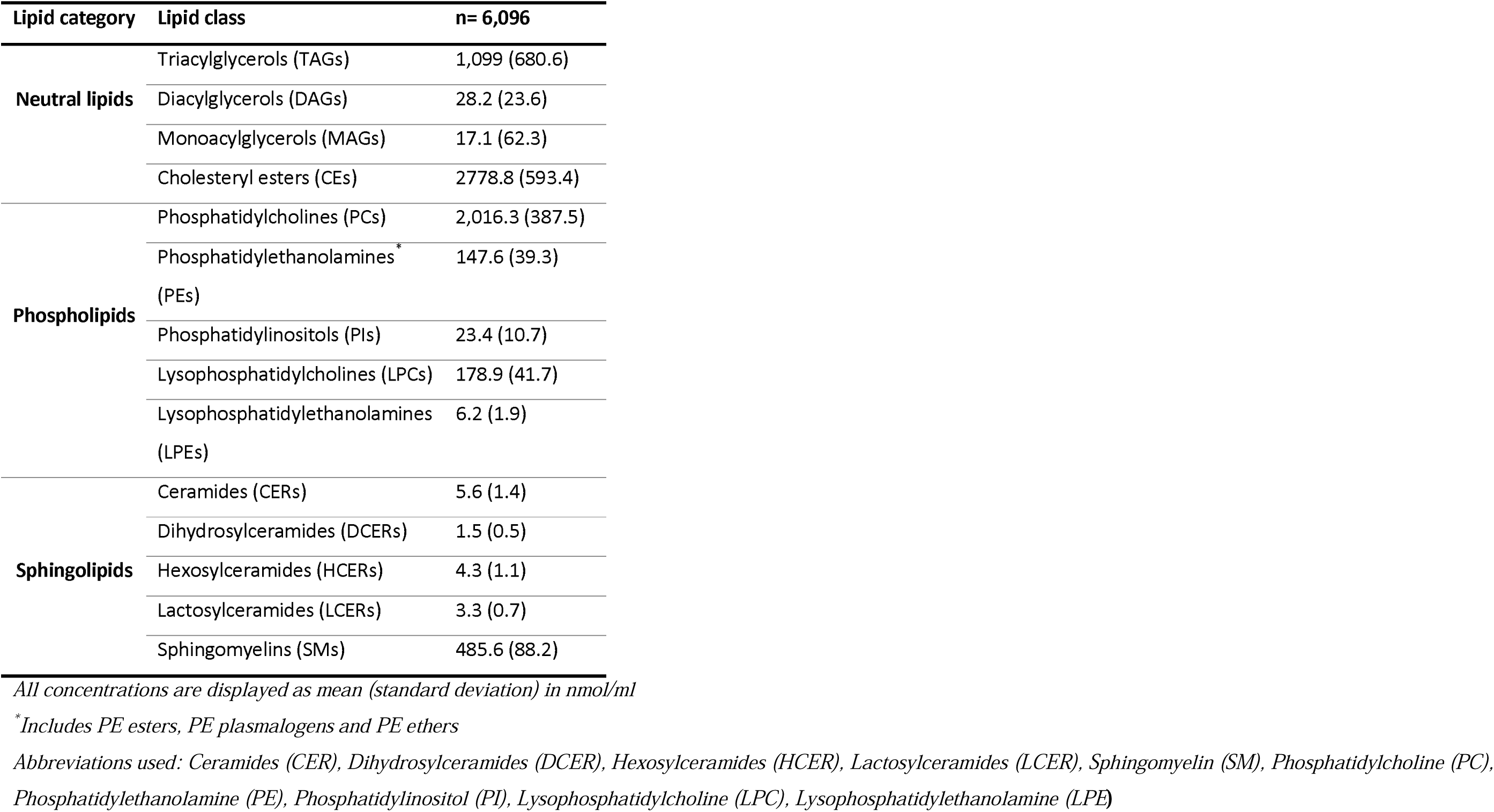
Overview of the total lipid concentrations per lipid class.

**Extended Table 2.** Overview of the FinnGen and EPIC-Potsdam sample characteristics.

| Characteristic | FinnGen<br>(n= 7,266) | EPIC-Potsdam<br>participants<br>(n= 1,188 ) |
| --- | --- | --- |
| Age (years) | 55.8 (5.8) | 50.3 (9.0) |
| Sex (women) | 4,642 (63.9) | 716 (60) |
| LDL-C (mg/dL) | 129.93 (35.96) | 122.0 (35.2) |
| HDL-C (mg/dL) | 64.20 (18.56) | 55.8 (14.7) |
| Triglycerides (mg/dL) | 112.48 (74.40) | 135.0 (100.0) |
| Cholesterol (mg/dL) | 215.78 (39.83) | 204.0 (42.4) |
| Use of lipid-lowering<br>medication (yes) | 805 (11.1) | 68 (5.7) |

*Data displayed represent the number of participants (percentages) for categorical variables and the mean (standard deviation) for continuous variables. Abbreviations: HDL-C = high-density lipoprotein cholesterol, LDL-C = low-density lipoprotein cholesterol.*

**Extended Table 3:** An overview of the lipid nomenclature and number of categories, classes, species and fatty acid composite measures.

| Lipid categories (n=3) | Lipid classes (n= 14) | Species (n= 970)* |  | Fatty acid composite (n= 267)* |  |
| --- | --- | --- | --- | --- | --- |
|  |  | Number (%) | Example | Number (%) | Example |
| Neutral lipids<br>(n= 628) | Triacylglycerols (TAGs) | 518 (53.1%) | TAG58:6(FA22:4)) | 21 (7.8%) | TAG(FA12:0) |
|  | Diacylglycerols (DAGs) | 58 (5.9%) | DAG(14:0/14:0) | 19 (7.0%) | DAG(FA18:3) |
|  | Monoacylglycerols (MAGs) | 26 (2.7%) | MAG(20:1) | Same as species |  |
|  | Cholesteryl esters (CEs) | 26 (2.7%) | CE(24:1) | Same as species |  |
| Phospholipids<br>(n= 286) | Phosphatidylcholines (PCs) | 104 (10.7%) | PC(16:0/20:3) | 23 (8.5%) | PC(FA20:1) |
|  | Phosphatidylethanolamines (PEs)** | 122 (127.5%) | PE(18:1/18:2) | 18 (7.0%) | PE(FA17:0) |
|  | Phosphatidylinositols (PIs) | 22 (2.7%) | PI(16:0/20:3) | 12 (4.8%) | PI(FA22:6) |
|  | Lysophosphatidylcholines (LPCs) | 18 (1.8%) | LPC(20:0) | Same as species |  |
|  | Lysophosphatidylethanolamines (LPEs) | 15 (1.6%) | LPE(15:0) | Same as species |  |
| Sphingolipids<br>(n= 61) | Sphingomyelins (SMs) | 12 (1.2%) | SM(18:1) | Same as species |  |
|  | Ceramides (CERs) | 12 (1.2%) | CER(22:1) | Same as species |  |
|  | Dihydrosylceramides (DCERs) | 13 (1.3%) | DCER(24:0) | Same as species |  |
|  | Hexosylceramides (HCERs) | 12 (1.2%) | HCER(26:0) | Same as species |  |
|  | Lactosylceramides (LCERs) | 12 (1.2%) | LCER(14:0) | Same as species |  |

\*Lipid species are defined by complete information on the total number of carbons and double bonds, whereas fatty acid composite measures sum up the concentration of all lipids within a class of a specific length and degree of saturation. In case of one-tailed lipids, the fatty acid composite measure is thus equal to the species. In addition, we also have information on the total fatty acid composition (n= 28), which sums up the concentration of a specific fatty acid tail across all lipid classes.
\*\*Containing PE esters, PE ethers, and PE plasmalogens

**Extended Table 4:**
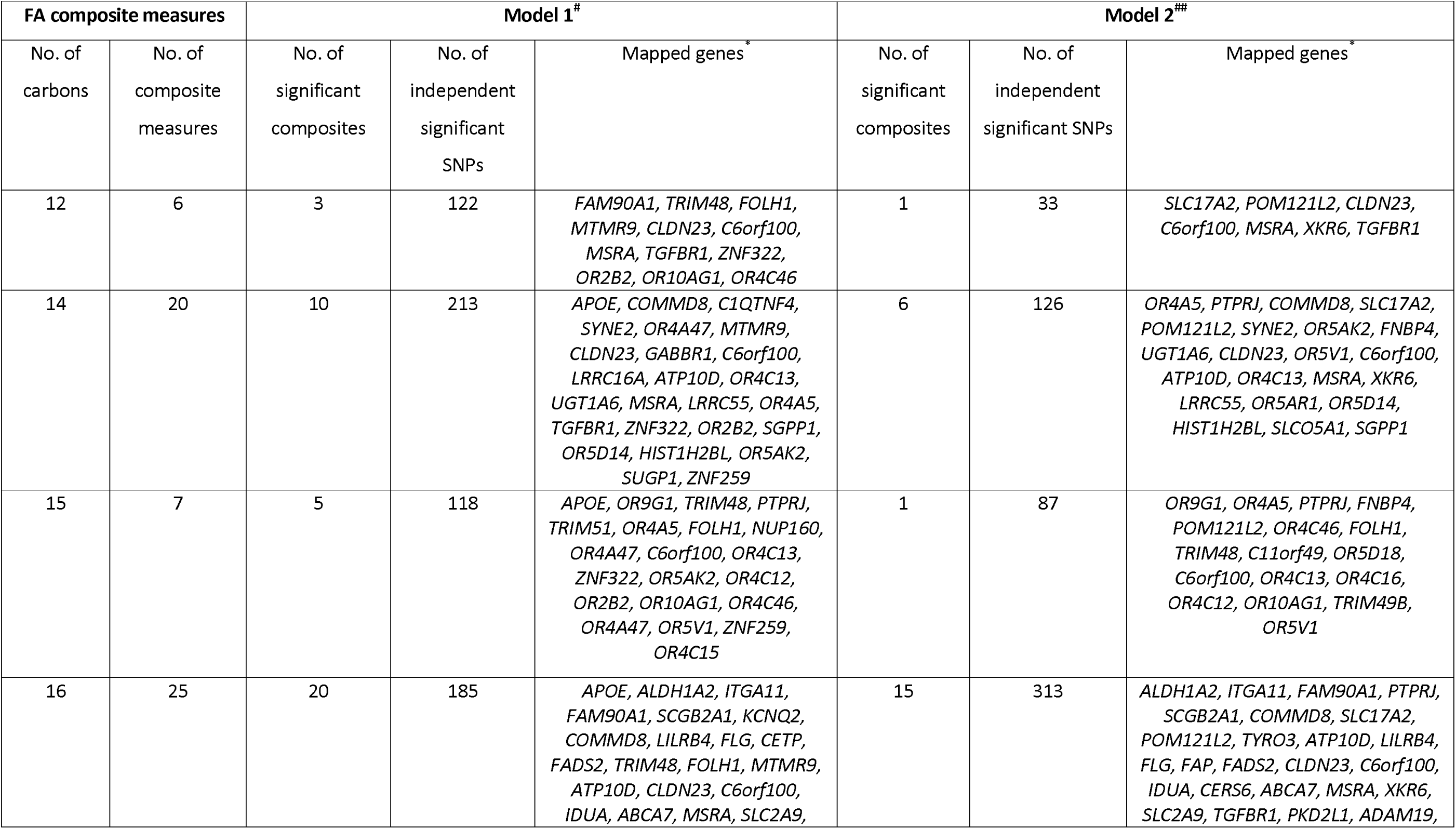
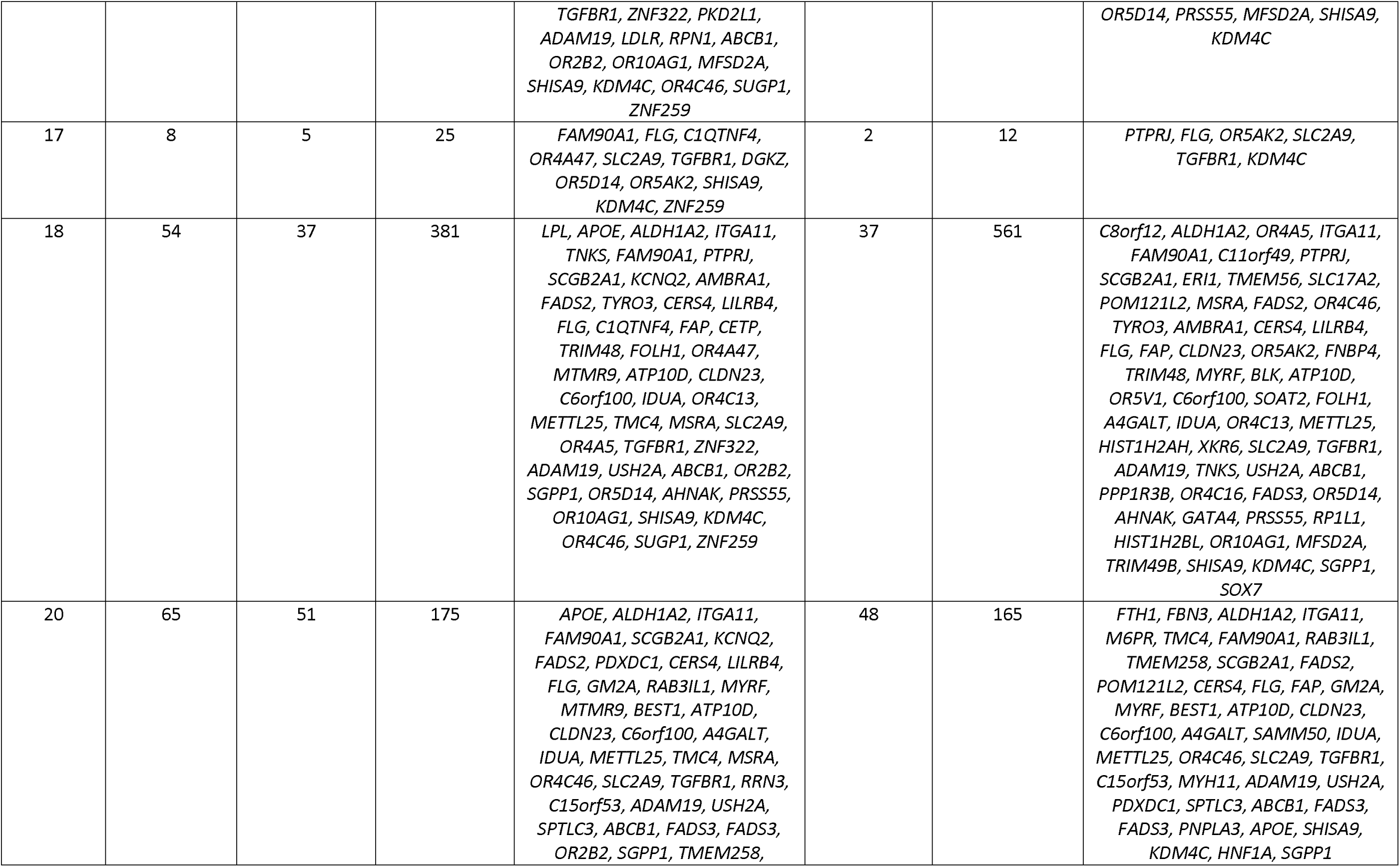
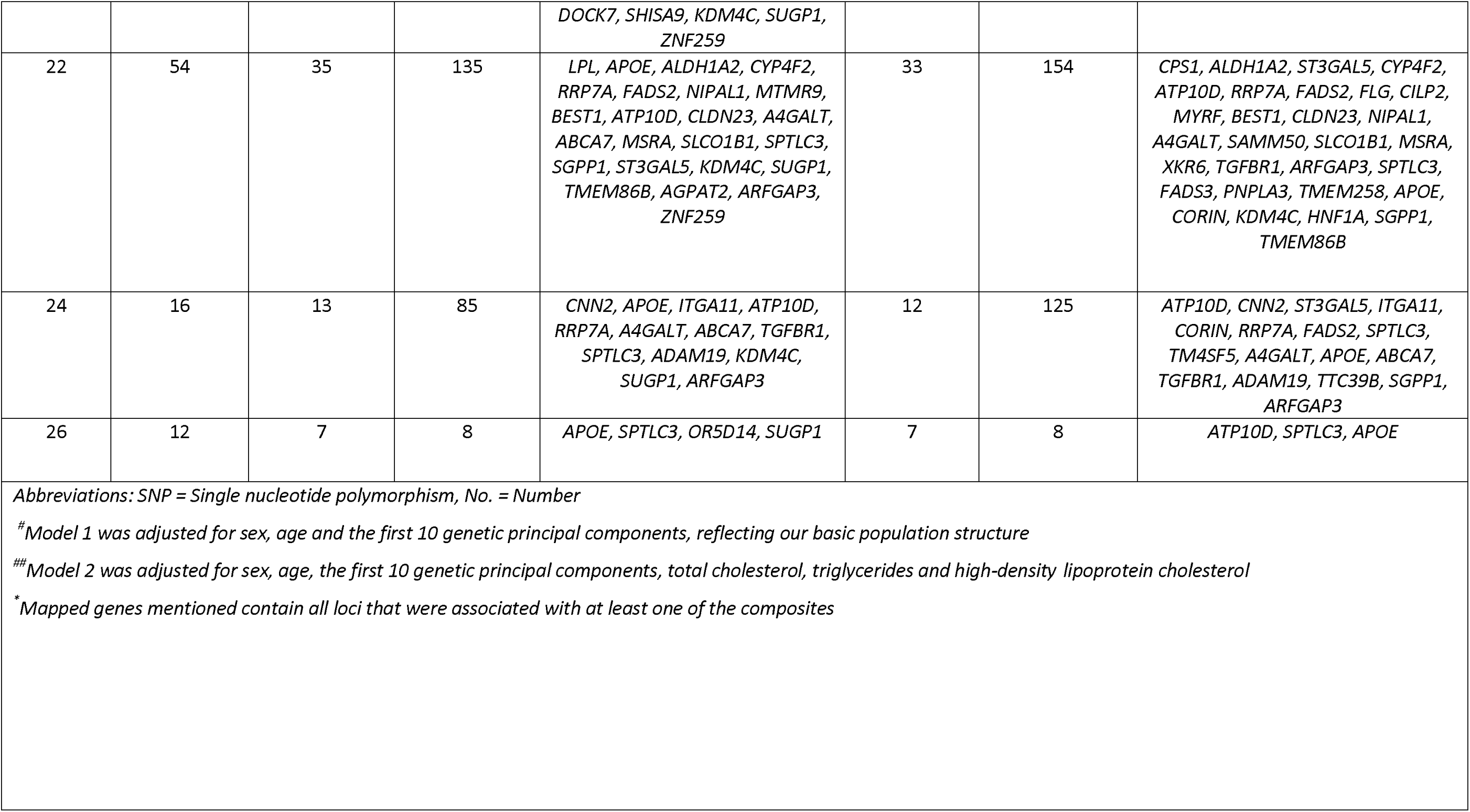
Overview of the number of metabolome-wide significant independent SNPs and genomic loci associated with fatty acid (FA) composite measures (n= 267) by the number of carbons.

|  |  |  |  |  |  |  |  |
| --- | --- | --- | --- | --- | --- | --- | --- |
| **FA composite measures** |  | **Model 1<sup>#</sup>** |  |  | **Model 2<sup>##</sup>** |  |  |
| No. of carbons | No. of composite measures | No. of significant composites | No. of independent significant SNPs | Mapped genes<sup>*</sup> | No. of significant composites | No. of independent significant SNPs | Mapped genes<sup>*</sup> |
| 12 | 6 | 3 | 122 | *FAM90A1, TRIM48, FOLH1, MTMR9, CLDN23, C6orf100, MSRA, TGFB1, ZNF322, OR2B2, OR10AG1, OR4C46* | 1 | 33 | *SLC17A2, POM121L2, CLDN23, C6orf100, MSRA, XKR6, TGFB1* |
| 14 | 20 | 10 | 213 | *APOE, COMMD8, C1QTNF4, SYNE2, OR4A47, MTMR9, CLDN23, GABBR1, C6orf100, LRRC16A, ATP10D, OR4C13, UGT1A6, MSRA, LRRC55, OR4A5, TGFB1, ZNF322, OR2B2, SGPP1, OR5D14, HIST1H2BL, OR5AK2, SUGP1, ZNF259* | 6 | 126 | *OR4A5, PTPRJ, COMMD8, SLC17A2, POM121L2, SYNE2, OR5AK2, FNBP4, UGT1A6, CLDN23, OR5V1, C6orf100, ATP10D, OR4C13, MSRA, XKR6, LRRC55, OR5AR1, OR5D14, HIST1H2BL, SLC05A1, SGPP1* |
| 15 | 7 | 5 | 118 | *APOE, OR9G1, TRIM48, PTPRJ, TRIM51, OR4A5, FOLH1, NUP160, OR4A47, C6orf100, OR4C13, ZNF322, OR5AK2, OR4C12, OR2B2, OR10AG1, OR4C46, OR4A47, OR5V1, ZNF259, OR4C15* | 1 | 87 | *OR9G1, OR4A5, PTPRJ, FNBP4, POM121L2, OR4C46, FOLH1, TRIM48, C11orf49, OR5D18, C6orf100, OR4C13, OR4C16, OR4C12, OR10AG1, TRIM49B, OR5V1* |
| 16 | 25 | 20 | 185 | *APOE, ALDH1A2, ITGA11, FAM90A1, SCGB2A1, KCNQ2, COMMD8, LILRB4, FLG, CETP, FADS2, TRIM48, FOLH1, MTMR9, ATP10D, CLDN23, C6orf100, IDUA, ABCA7, MSRA, SLC2A9,* | 15 | 313 | *ALDH1A2, ITGA11, FAM90A1, PTPRJ, SCGB2A1, COMMD8, SLC17A2, POM121L2, TYRO3, ATP10D, LILRB4, FLG, FAP, FADS2, CLDN23, C6orf100, IDUA, CERS6, ABCA7, MSRA, XKR6, SLC2A9, TGFB1, PKD2L1, ADAM19,* |
|  |  |  |  | TGFB1, ZNF322, PKD2L1, ADAM19, LDLR, RPN1, ABCB1, OR2B2, OR10AG1, MFSD2A, SHISA9, KDM4C, OR4C46, SUGP1, ZNF259 |  |  | OR5D14, PRSS55, MFSD2A, SHISA9, KDM4C |
| 17 | 8 | 5 | 25 | FAM90A1, FLG, C1QTNF4, OR4A47, SLC2A9, TGFB1, DGKZ, OR5D14, OR5AK2, SHISA9, KDM4C, ZNF259 | 2 | 12 | PTPRJ, FLG, OR5AK2, SLC2A9, TGFB1, KDM4C |
| 18 | 54 | 37 | 381 | LPL, APOE, ALDH1A2, ITGA11, TNKS, FAM90A1, PTPRJ, SCGB2A1, KCNQ2, AMBRA1, FADS2, TYRO3, CERS4, LILRB4, FLG, C1QTNF4, FAP, CETP, TRIM48, FOLH1, OR4A47, MTMR9, ATP10D, CLDN23, C6orf100, IDUA, OR4C13, METTL25, TMC4, MSRA, SLC2A9, OR4A5, TGFB1, ZNF322, ADAM19, USH2A, ABCB1, OR2B2, SGPP1, OR5D14, AHNK, PRSS55, OR10AG1, SHISA9, KDM4C, OR4C46, SUGP1, ZNF259 | 37 | 561 | C8orf12, ALDH1A2, OR4A5, ITGA11, FAM90A1, C11orf49, PTPRJ, SCGB2A1, ERI1, TMEM56, SLC17A2, POM121L2, MSRA, FADS2, OR4C46, TYRO3, AMBRA1, CERS4, LILRB4, FLG, FAP, CLDN23, OR5AK2, FBNP4, TRIM48, MYRF, BLK, ATP10D, OR5V1, C6orf100, SOAT2, FOLH1, A4GALT, IDUA, OR4C13, METTL25, HIST1H2AH, XKR6, SLC2A9, TGFB1, ADAM19, TNKS, USH2A, ABCB1, PPP1R3B, OR4C16, FADS3, OR5D14, AHNK, GATA4, PRSS55, RP11L1, HIST1H2BL, OR10AG1, MFSD2A, TRIM49B, SHISA9, KDM4C, SGPP1, SOX7 |
| 20 | 65 | 51 | 175 | APOE, ALDH1A2, ITGA11, FAM90A1, SCGB2A1, KCNQ2, FADS2, PDXDC1, CERS4, LILRB4, FLG, GM2A, RAB31L1, MYRF, MTMR9, BEST1, ATP10D, CLDN23, C6orf100, A4GALT, IDUA, METTL25, TMC4, MSRA, OR4C46, SLC2A9, TGFB1, RRN3, C15orf53, ADAM19, USH2A, SPTLC3, ABCB1, FADS3, FADS3, OR2B2, SGPP1, TMEM258, | 48 | 165 | FTH1, FBN3, ALDH1A2, ITGA11, M6PR, TMC4, FAM90A1, RAB31L1, TMEM258, SCGB2A1, FADS2, POM121L2, CERS4, FLG, FAP, GM2A, MYRF, BEST1, ATP10D, CLDN23, C6orf100, A4GALT, SAMM50, IDUA, METTL25, OR4C46, SLC2A9, TGFB1, C15orf53, MYH11, ADAM19, USH2A, PDXDC1, SPTLC3, ABCB1, FADS3, FADS3, PNPLA3, APOE, SHISA9, KDM4C, HNF1A, SGPP1 |
|  |  |  |  | *DOCK7, SHISA9, KDM4C, SUGP1, ZNF259* |  |  |  |
| 22 | 54 | 35 | 135 | *LPL, APOE, ALDH1A2, CYP4F2, RRP7A, FADS2, NIPAL1, MTMR9, BEST1, ATP10D, CLDN23, A4GALT, ABCA7, MSRA, SLCO1B1, SPTLC3, SGPP1, ST3GAL5, KDM4C, SUGP1, TMEM86B, AGPAT2, ARFGAP3, ZNF259* | 33 | 154 | *CPS1, ALDH1A2, ST3GAL5, CYP4F2, ATP10D, RRP7A, FADS2, FLG, CILP2, MYRF, BEST1, CLDN23, NIPAL1, A4GALT, SAMM50, SLCO1B1, MSRA, XKR6, TGFB1, ARFGAP3, SPTLC3, FADS3, PNPLA3, TMEM258, APOE, CORIN, KDM4C, HNF1A, SGPP1, TMEM86B* |
| 24 | 16 | 13 | 85 | *CNN2, APOE, ITGA11, ATP10D, RRP7A, A4GALT, ABCA7, TGFB1, SPTLC3, ADAM19, KDM4C, SUGP1, ARFGAP3* | 12 | 125 | *ATP10D, CNN2, ST3GAL5, ITGA11, CORIN, RRP7A, FADS2, SPTLC3, TM4SF5, A4GALT, APOE, ABCA7, TGFB1, ADAM19, TTC39B, SGPP1, ARFGAP3* |
| 26 | 12 | 7 | 8 | *APOE, SPTLC3, OR5D14, SUGP1* | 7 | 8 | *ATP10D, SPTLC3, APOE* |

*Abbreviations: SNP = Single nucleotide polymorphism, No. = Number*
*# Model 1 was adjusted for sex, age and the first 10 genetic principal components, reflecting our basic population structure*
*## Model 2 was adjusted for sex, age, the first 10 genetic principal components, total cholesterol, triglycerides and high-density lipoprotein cholesterol*
*\* Mapped genes mentioned contain all loci that were associated with at least one of the composites*

**Extended Table 5:**
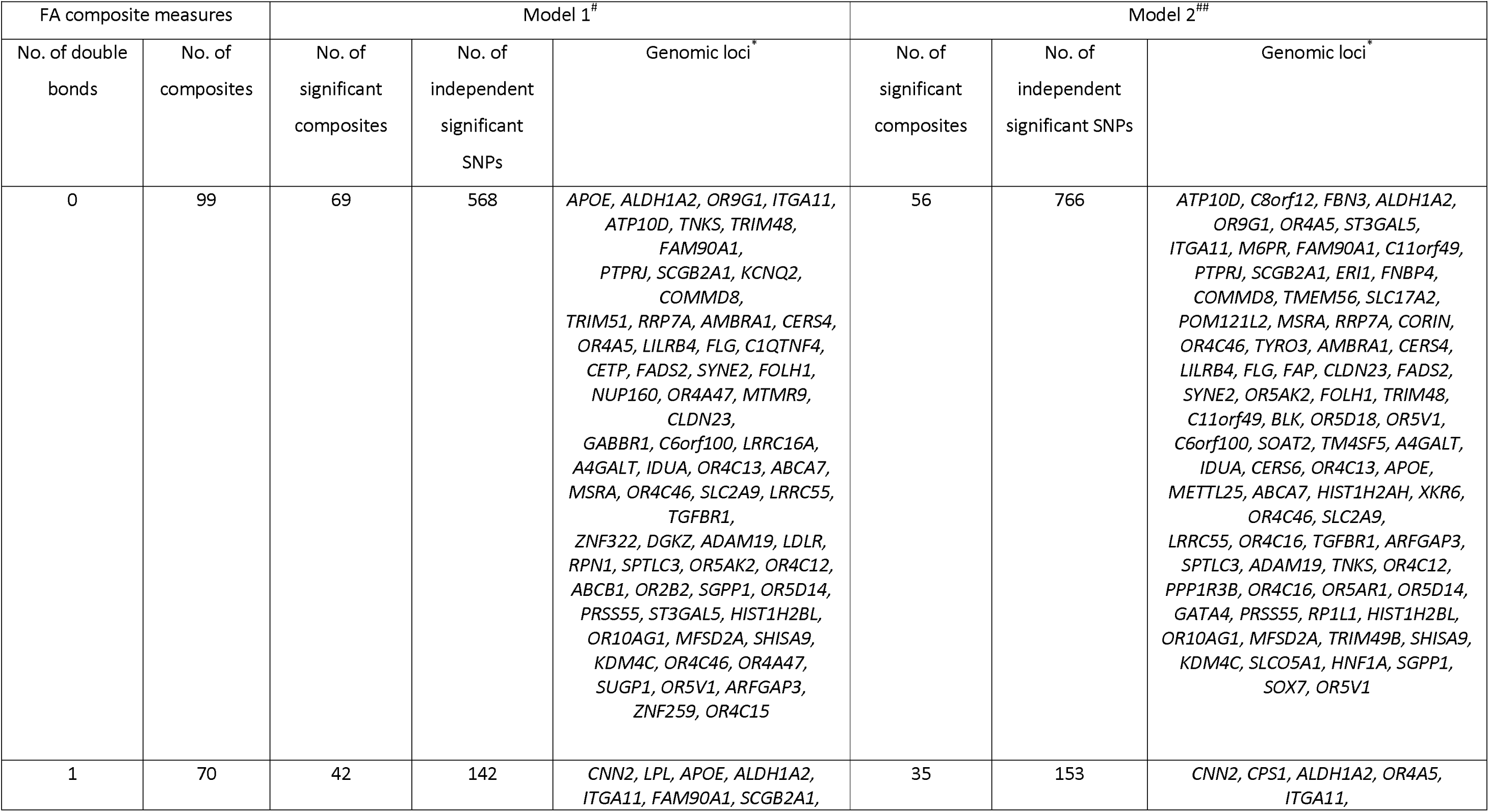
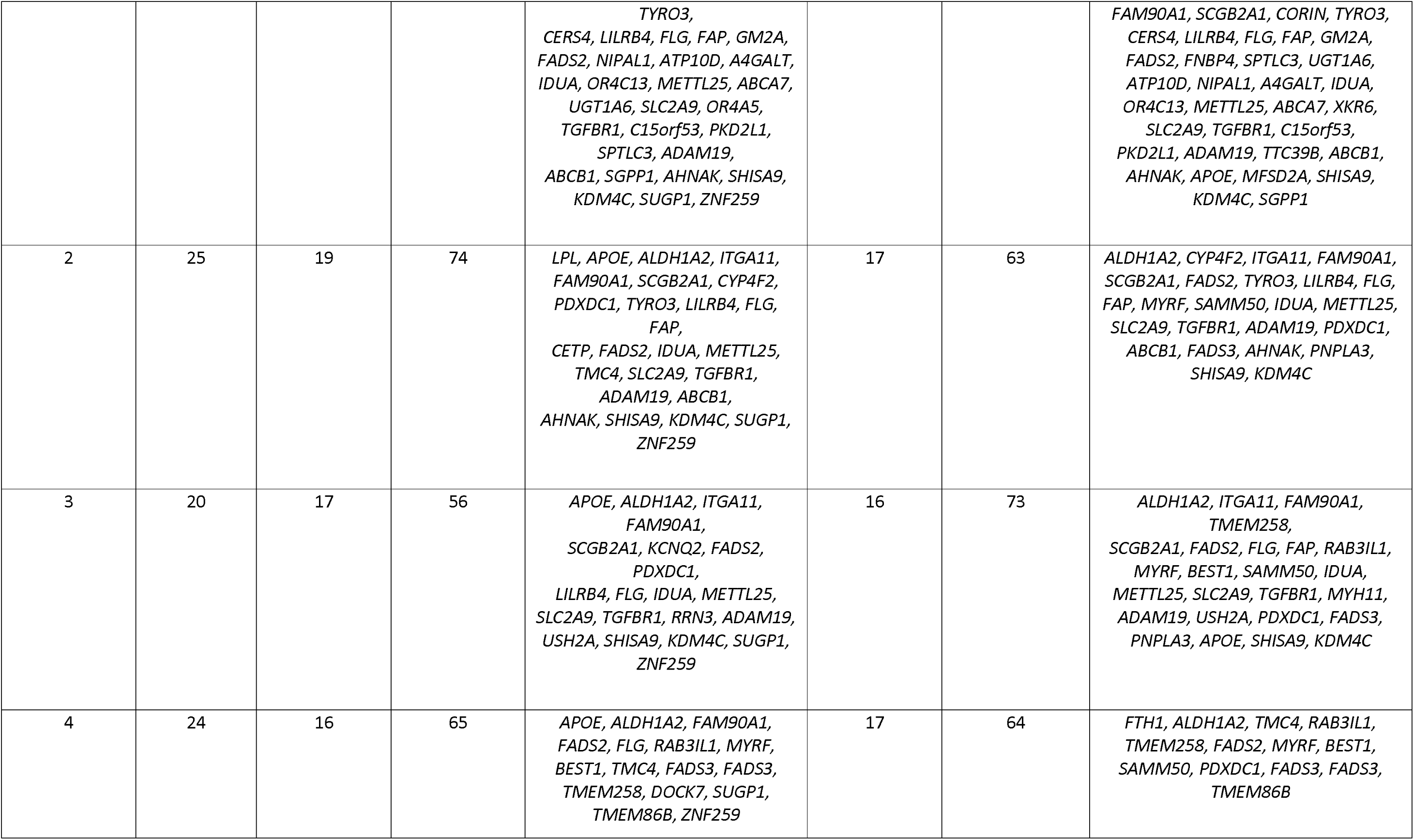
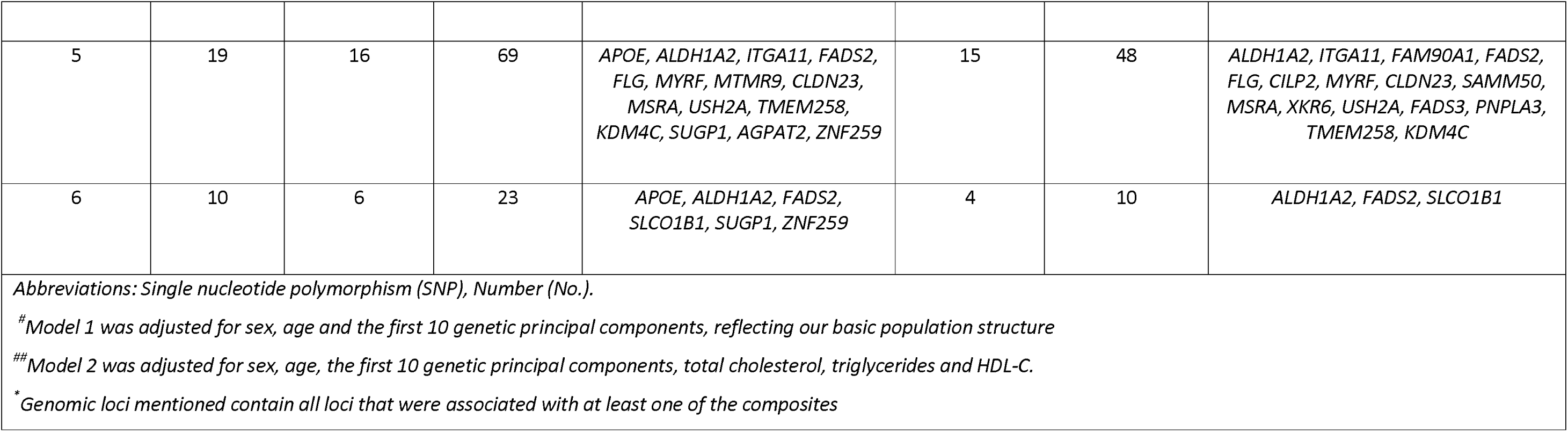
Overview of the number of metabolome-wide significant independent SNPs and genomic loci with fatty acid (FA) composite measures (n= 267) by the number of double bonds.

**Extended Table 6:** GWAS replication results per lipid class in the EPIC-Potsdam cohort.

| Lipid class | No. of lipid species in Rhineland Study | Model 1 |  |  | Model 2 |  |  |
| --- | --- | --- | --- | --- | --- | --- | --- |
|  |  | Metabolome-wide significant lipid species in Rhineland Study | Replicated number of species per lipid class in EPIC-Potsdam | Replication (%) per lipid class | Metabolome-wide significant lipid species in Rhineland Study | Replicated number of species per lipid class in EPIC-Potsdam | Replication (%) per lipid class |
| TAG | 518 | 405 | 383 | 94.6 | 230 | 178 | 77.4 |
| PE | 58 | 24 | 22 | 91.7 | 25 | 22 | 88.0 |
| PEP | 44 | 12 | 11 | 91.7 | 11 | 11 | 100 |
| PC | 104 | 41 | 37 | 90.2 | 44 | 41 | 93.2 |
| DAG | 58 | 44 | 39 | 88.6 | 34 | 27 | 79.4 |
| LPC | 18 | 8 | 6 | 75.0 | 11 | 8 | 72.7 |
| HCER | 11 | 9 | 7 | 77.8 | 10 | 7 | 70.0 |
| LCER | 12 | 8 | 6 | 75.0 | 10 | 8 | 80.0 |
| PI | 22 | 15 | 11 | 73.3 | 15 | 14 | 93.3 |
| CER | 12 | 7 | 5 | 71.4 | 8 | 4 | 50 |
| MAG | 26 | 9 | 6 | 66.7 | 9 | 5 | 55.6 |
| CE | 26 | 21 | 13 | 61.9 | 17 | 14 | 82.3 |
| SM | 12 | 12 | 7 | 58.3 | 10 | 7 | 70.0 |
| PEO | 20 | 6 | 3 | 50 | 6 | 4 | 66.7 |
| LPE | 15 | 9 | 5 | 55.5 | 9 | 5 | 55.5 |
| DCER | 13 | 8 | 4 | 50.0 | 7 | 6 | 85.7 |

**Extended Figure 1:**
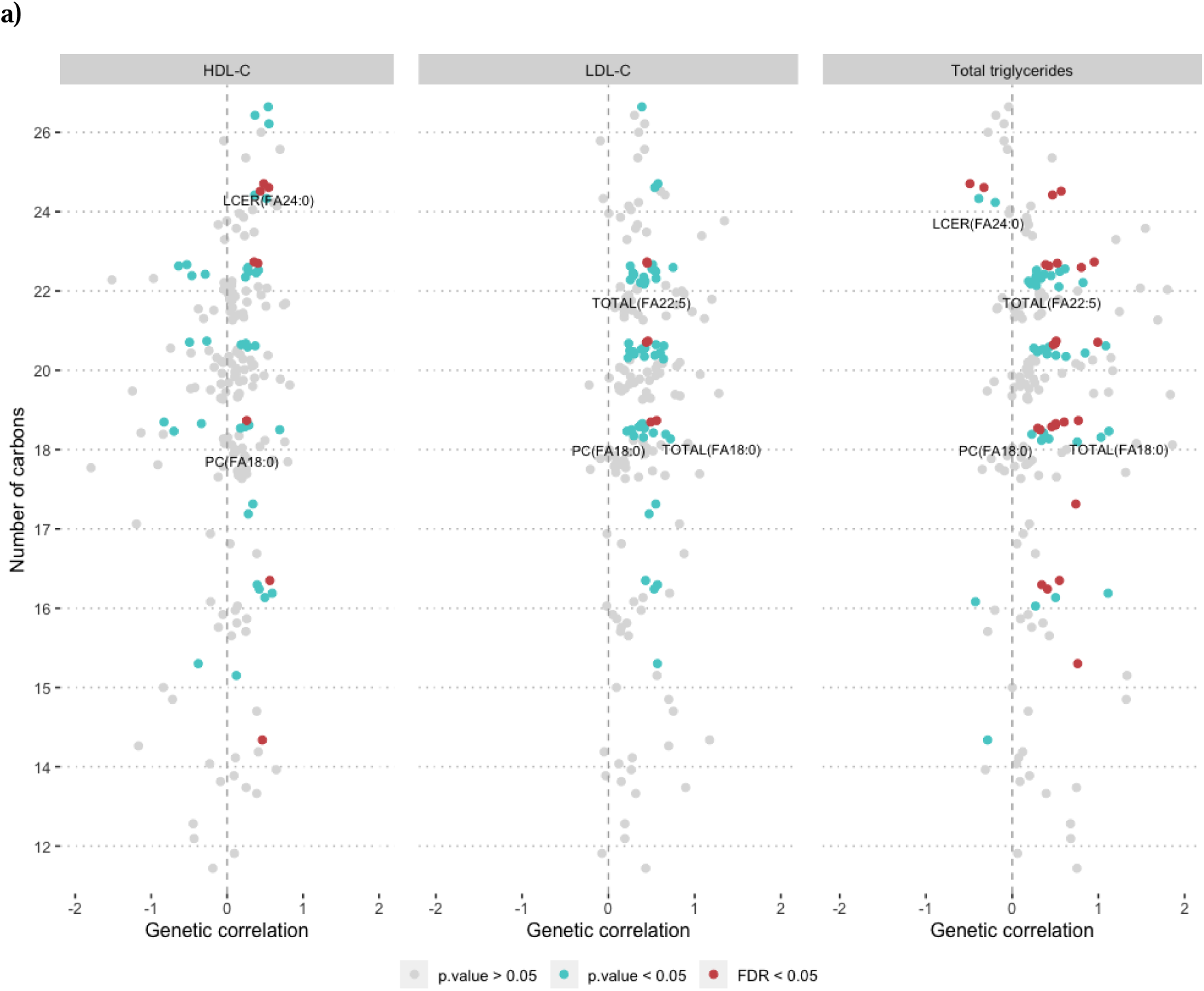
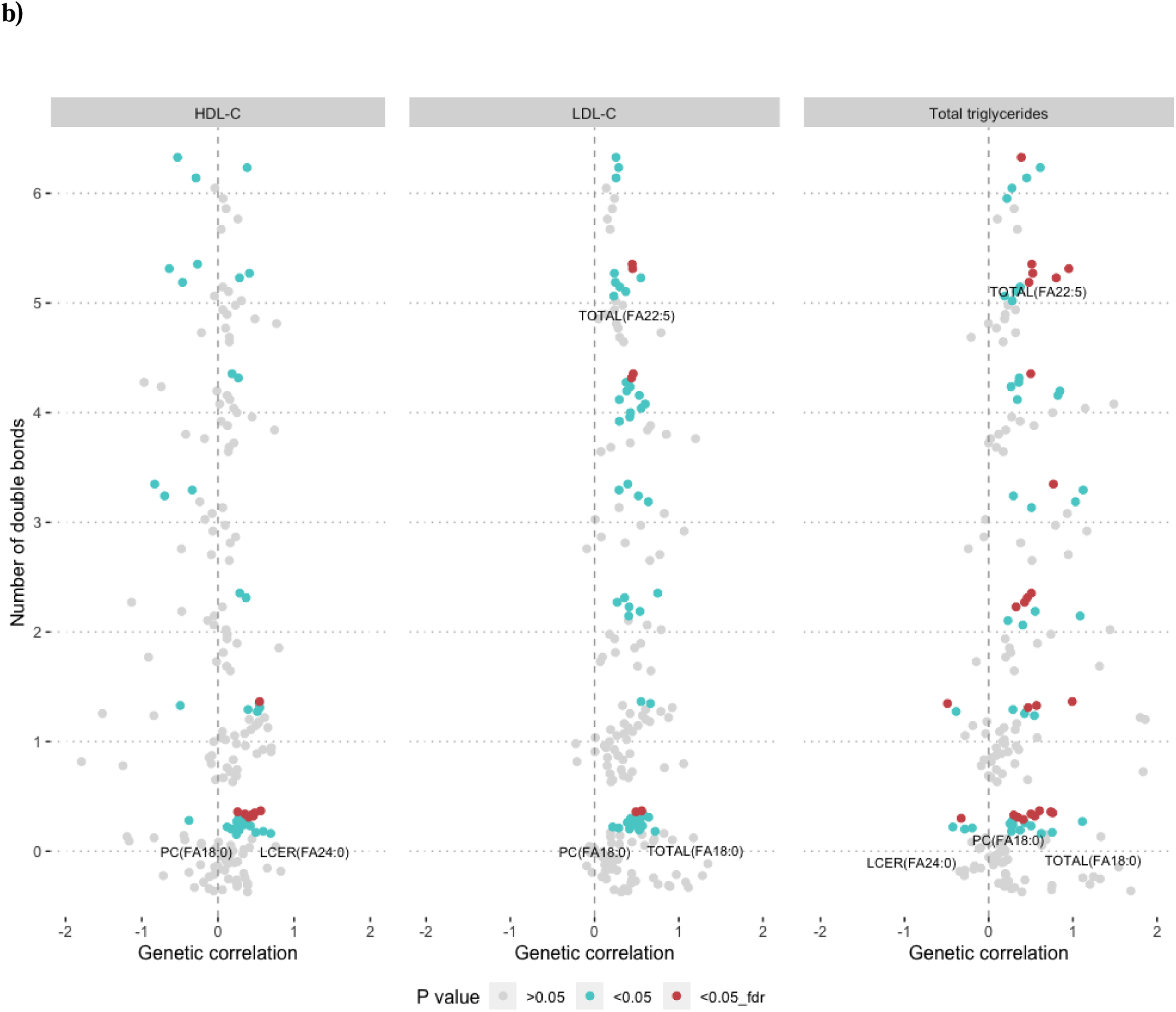
Genetic correlation of fatty acid composite measures with traditional lipid measurements. The genetic correlation of fatty acid composite measures by the number of carbons **(a)**, or double bonds **(b)**, with HDL-C, LDL-C and total triglycerides after adjustment for age, sex and the first 10 genetic principal components. Statistical significance is indicated by colours. Fatty acid composite measures based on genetic correlation within each class, which overlapped with at least two clinical lipid measurements, are annotated.

**Extended Figure 2:**
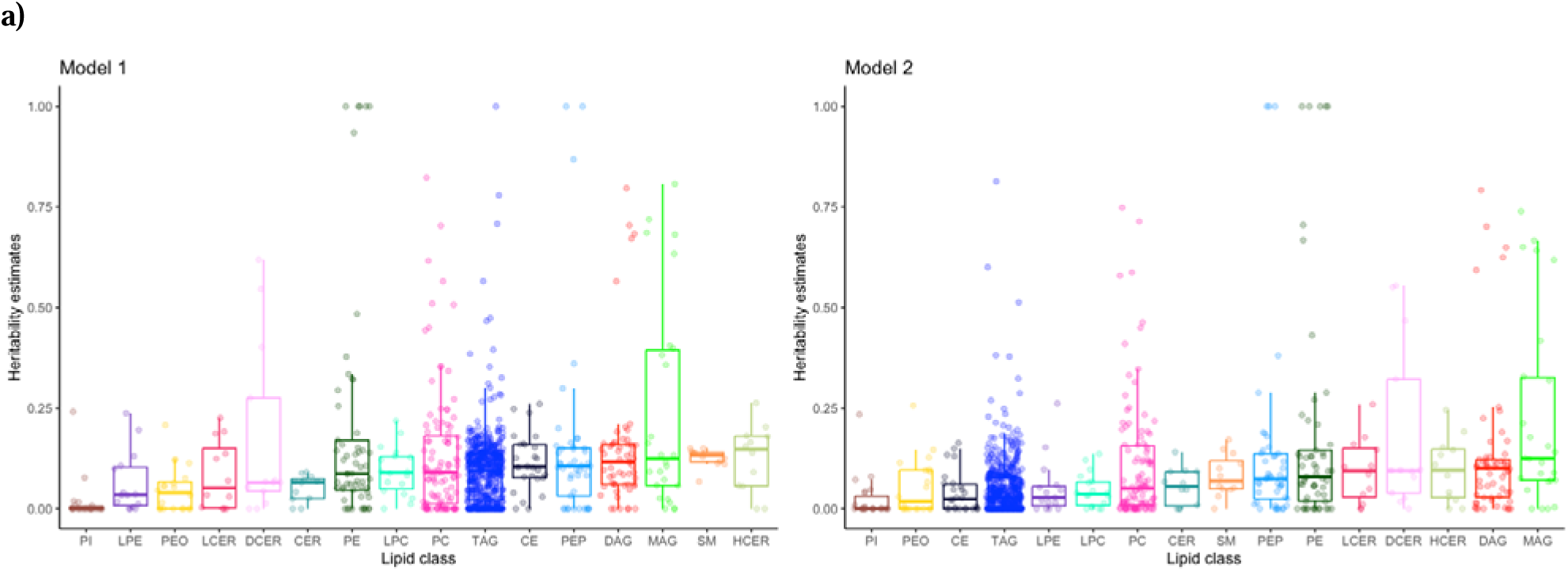
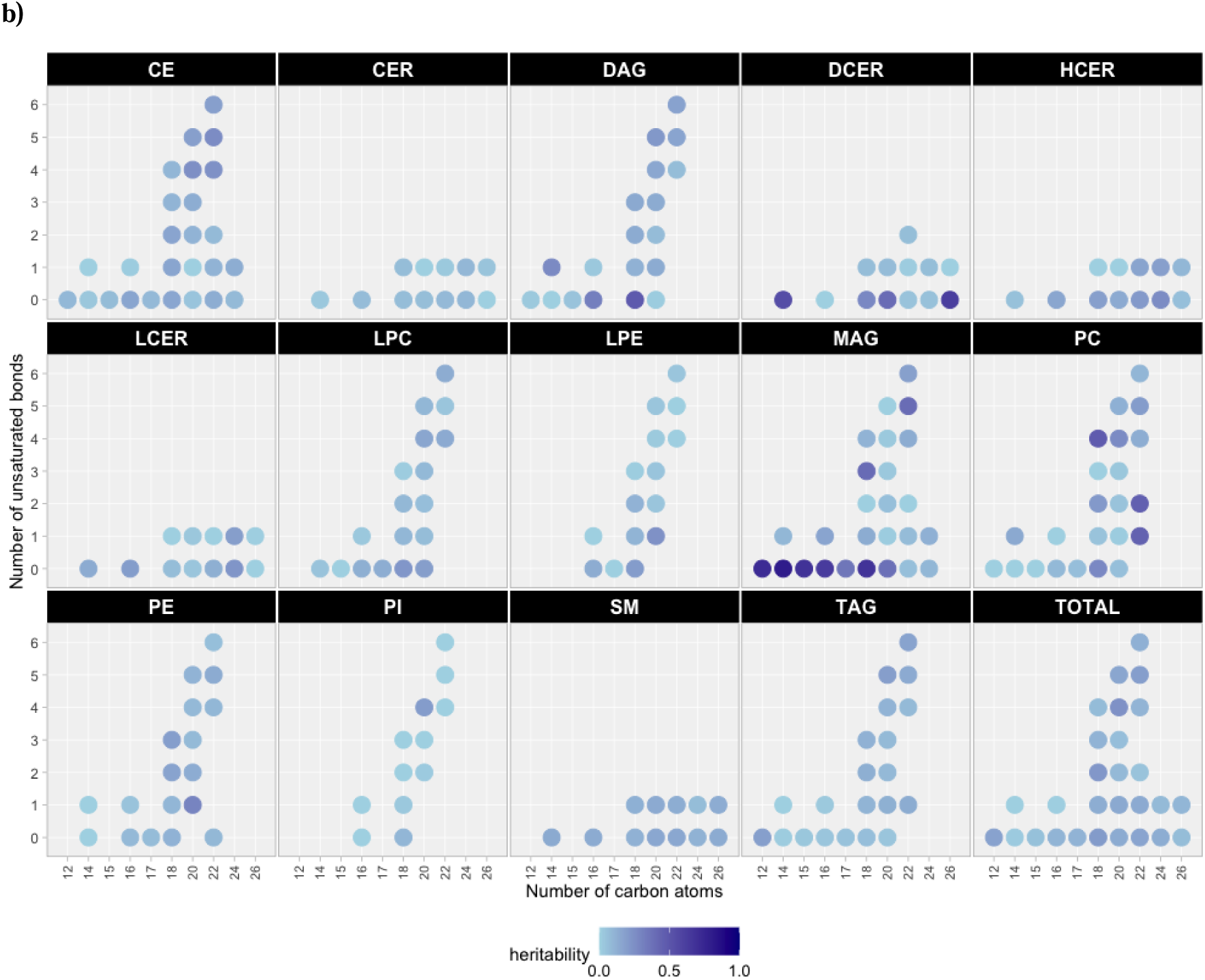
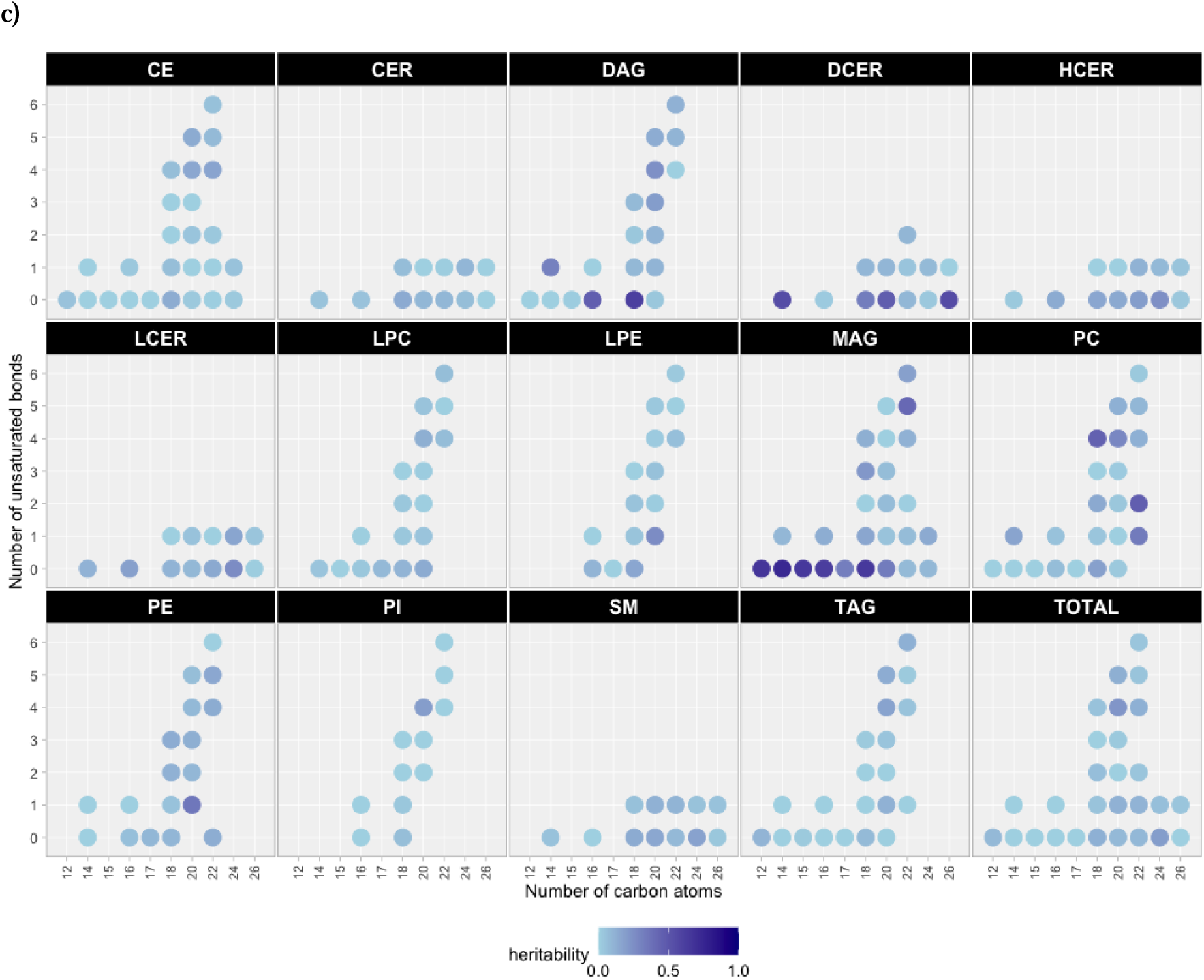
Heritability estimates for lipid species and fatty acid composition. **(a)** Boxplots display the different SNP heritability estimates based on Genome-wide Complex Trait Analysis (GCTA) for model 1 (adjusted for age, sex and the first 10 genetic principal components), and model 2 (additionally adjusted for LDL-C, HDL-C, triglycerides and use of lipid-lowering medication) for lipid species within respective classes. The lipid classes are sorted from left to right based on the median value of the heritability estimates of their lipid species. The degree of heritability for fatty acid composition by the number of carbon atoms and unsaturated bonds for model 1 **(b)**, and model 2 **(c)**, with lighter colour and larger disk size indicating a higher heritability estimate.

**Extended Figure 3:**
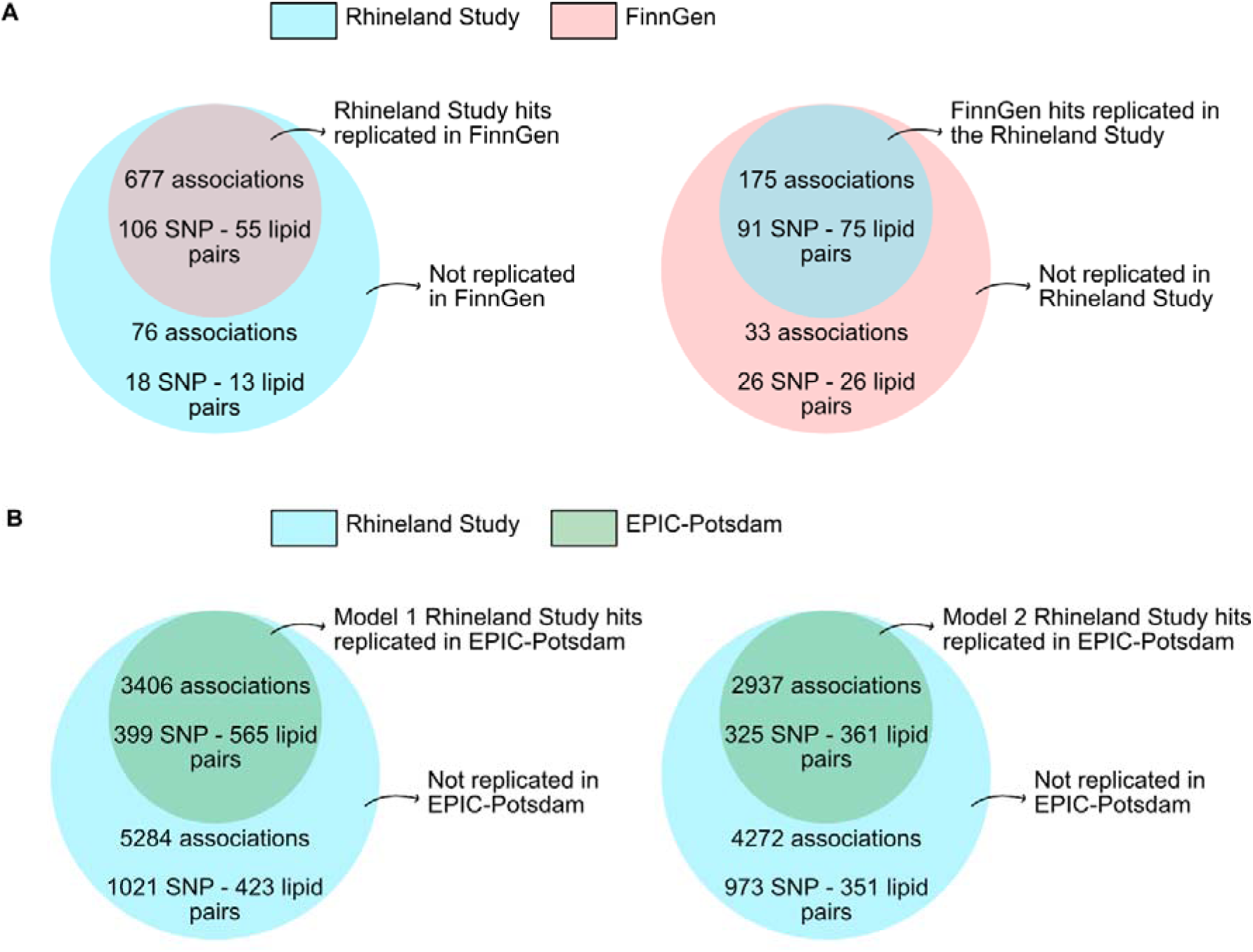
GWAS replication results. **(a)** The metabolome-wide significant SNP-lipid pair associations overlapping between the Rhineland Study and the FinnGen cohort for model 1 (adjusted for age, sex, and the first 10 genetic principal components), with replication defined as the association reaching nominal significance in the other cohort. **(b)** The metabolome-wide significant SNP-lipid pair associations overlapping between the Rhineland Study and the EPIC-Potsdam cohort for model 1 (adjusted for age, sex, and the first 10 genetic principal components) and model 2 (additionally adjusted for LDL-C, HDL-C, triglycerides and use of lipid-lowering medication), with replication defined as the association reaching nominal significance in the EPIC-Potsdam cohort.. *Abbreviations: HDL-C = high-density lipoprotein cholesterol, LDL-C = low-density lipoprotein cholesterol*.

